# Genome-wide detection and clinical prioritization of tandem repeat outliers using long-read sequencing

**DOI:** 10.64898/2026.04.30.26352103

**Authors:** Sophia B. Gibson, Nikhita Damaraju, J. Gus Gustafson, Elsa V. Balton, Sirisak Chanprasert, Ian A. Glass, Martha Horike-Pyne, Runjun D. Kumar, Kathleen A. Leppig, Chris Lundberg, Jane Ranchalis, Elisabeth A. Rosenthal, Andrew K. Solomon, Andrew B. Stergachis, Mark Wener, Undiagnosed Diseases Network, Gail P. Jarvik, Elizabeth E Blue, Katrina M. Dipple, Harriet Dashnow, Lea M. Starita, Danny E. Miller

**Author notes:** **Contact:** Danny E. Miller.

## Abstract

**Background:** Tandem repeat expansions (TREs) cause over 60 known neurological, neuromuscular, and developmental disorders. Detecting these expansions genome-wide is challenging due to their size, sequence complexity (including interruptions), and population variation. While long-read sequencing is an emerging technology that can fully resolve many TREs, no methods have been described for genome-wide identification and prioritization of candidate pathogenic TREs with this technology.

**Methods:** Using a newly developed pipeline called TRoLR (Tandem Repeat outliers identified with Long Reads), we analyzed haplotype-resolved long-read genome assemblies from 471 ancestrally diverse individuals to define population distributions for over three million tandem repeat loci, capturing clinically relevant interruptions. Outlier expansions were identified relative to these distributions and prioritized by genomic location and comparison to known pathogenic loci. The framework was applied to 47 cases from the Undiagnosed Diseases Network.

**Results:** Population stratification of repeat metrics was observed at 7% of loci, with highest variability among individuals of African ancestry. Outlier analysis confirmed known pathogenic *CNBP* and *ATXN8OS* expansions, detected carrier-range alleles at *RFC1*, *CSTB*, and *FXN*, and revealed a novel CGG expansion in the 5’ UTR of *PCMTD2* exhibiting hypermethylation and intergenerational instability. Genome-wide screening also identified intronic pentanucleotide expansions at *IQCB1* and *MAP3K15* in controls composed of motifs that have been associated with pathogenicity at other disease loci.

**Conclusions:** Quantifying the longest uninterrupted repeat segment in long-read assemblies enables detection of clinically relevant repeat expansions and loss of stabilizing interruptions. This approach enhances both diagnostic confirmation and discovery of candidate pathogenic expansions, with implications for clinical interpretation and research into complex repeat-mediated disorders.

## INTRODUCTION

More than half of individuals with a suspected genetic disorder remain undiagnosed after comprehensive genetic testing^1^. A major contributor to this gap is the limited ability of short-read sequencing (SRS) to detect tandem repeat expansions (TREs). Tandem repeats are sequences of short DNA motifs repeated in direct, head-to-tail succession and are among the most polymorphic and mutationally dynamic elements in the human genome^2,3^. They are conventionally subdivided by motif length into short tandem repeats (STRs; motifs of 1–6 bp) and variable-number tandem repeats (VNTRs; motifs >6 bp), though this boundary is somewhat arbitrary and both classes exhibit high mutation rates. Expansions of tandem repeats cause more than 60 human disorders, including Huntington’s disease, fragile X syndrome, myotonic dystrophy, and multiple spinocerebellar ataxias with pathogenic motifs ranging from trinucleotides to hexanucleotides and beyond^4,5^. While resources like the STRchive database have catalogued known pathogenic tandem repeat loci and their associated clinical thresholds, TREs remain systematically under-ascertained in standard clinical testing^6^. This is because most are not included in exome or genome sequencing analysis pipelines and many disease-associated loci have been discovered only recently, suggesting the full landscape of pathogenic repeat expansions is likely far from complete.

Specialized SRS-based tools have been developed to address some of these challenges. ExpansionHunter, STRetch and GangSTR support genotyping at predefined loci, while ExpansionHunter Denovo and STRling enable discovery of expansions at novel loci.^7–11^ However, alleles may be thousands of base pairs in length, which exceed typical read lengths produced by SRS technologies (<150 bp)^12^. These limitations can lead to both false negatives–where pathogenic expansions go undetected–and inaccurate genotyping of repeat length and motif composition^12^.

Long-read sequencing (LRS) technologies, including those from Oxford Nanopore Technologies (ONT) and Pacific Biosciences (PacBio), offer a solution. With read lengths routinely exceeding tens of kilobases, LRS reads can span most tandem repeat expansions, enabling direct measurement of repeat length and precise characterization of motif composition.^13,14^ The ability to generate haplotype-resolved genome assemblies from LRS data provides additional advantages, allowing evaluation of phased repeat alleles and heterozygosity. The ability to perform comprehensive and simultaneous evaluation of repeat length and motif composition is driving research and clinical interest in the use of LRS for evaluating individuals for disease-causing repeat expansions, with several groups demonstrating improved diagnostic yields compared to SRS-based approaches.^15–17^

However, the transition from LRS data generation to actionable variant calls for tandem repeats remains challenging. While numerous tools now exist for repeat genotyping from long reads (e.g., TRGT, Straglr, STRkit, and vamos), the integration of these calls with population-level reference data, genomic annotation, and clinical prioritization has required substantial manual effort^18–22^. Furthermore, most publicly available population allele frequency data for tandem repeats are from SRS studies, such as TRatlas (wlcb.oit.uci.edu/TRatlas), which are not able to accurately evaluate the true distribution of repeat lengths or motif sequences genome-wide.^12^ This mismatch between LRS data and SRS-derived reference databases limits the utility of existing annotation approaches.

Recent efforts have begun to address these gaps more broadly, such as work from the *All of Us* Research Program, which introduced the longest pure repeat segment (LPS) metric as a means of characterizing tandem repeat alleles.^23,24^ The LPS represents the longest uninterrupted run of a given motif within a repeat array and provides a more nuanced view of repeat structure than total repeat length alone. This metric is inspired by the observation that many pathogenic repeat expansions are characterized by the loss of stabilizing interruptions which may precede overt expansion and contribute to disease pathogenesis (e.g. CAA interruptions in the CAG tract to stabilize Huntington’s Disease)^25^.

Separately, population-scale LRS initiatives are generating diverse reference datasets needed to establish accurate population tandem repeat motif compositions and length distributions.^12,26–28^ Online resources, including TRExplorer (trexplorer.broadinstitute.org), enable interactive querying of population-level tandem repeat variation from long read data at scale^6,29,30^.

With the expanding availability of LRS data, there is growing power for leveraging it for tandem repeat outlier detection. This principle—identifying expanded alleles as statistical outliers relative to a control population—was first developed for short-read data by STRetch, ExpansionHunter Denovo, and STRling^8–10^. These tools demonstrated the utility of prioritizing both known and novel pathogenic expansions without prior specification of candidate loci. Translating this paradigm to LRS data has the potential to substantially improve both sensitivity and resolution, yet current LRS tools address only components of the problem.

For example, TRGT and TRVZ enable accurate genotyping and allele-level visualization from PacBio HiFi data but do not incorporate population reference distributions or automated outlier calling. PathSTR provides population-level visualization at medically relevant loci but is restricted to ∼70 known disease-associated repeats and does not perform genome-wide analysis. TREAT/otter supports outlier detection but requires users to supply their own control cohort, presenting a practical barrier for clinical use.^22^ The SUMMER pipeline integrates multiple variant classes from nanopore data but limits tandem repeat analysis to 55 known pathogenic loci. None of these tools combines genome-wide outlier detection from long-read assemblies with population comparison, genomic annotation, and interactive clinical reporting in a single automated workflow.

Despite this rapid progress in genotyping and population cataloging, no existing tool provides an end-to-end pipeline for genome-wide tandem repeat outlier detection from long-read data with integrated clinical prioritization. Here we present TRoLR (Tandem Repeat Outliers identified with Long Reads), a comprehensive pipeline that integrates these advances into a unified workflow for tandem repeat outlier detection **(Figure 1)**. TRoLR takes haplotype-aware genome assemblies as input, genotypes tandem repeats genome-wide using vamos, calculates the LPS at each locus, and identifies statistical outliers by comparison to population reference data derived from 471 individuals. Outliers are annotated with genomic context from GENCODE. The pipeline produces an interactive HTML report that organizes candidates by genomic context and clinical relevance, enabling rapid prioritization for research or clinical interpretation. We demonstrate the utility of TRoLR by first characterizing the LPS landscape across 471 diverse individuals from the 1000 Genomes Project, establishing population-level distributions at over 3 million loci. We then apply TRoLR to 47 individuals from the Undiagnosed Diseases Network (UDN) where we identify and prioritize previously known pathogenic expansions in *CNBP* and *ATXN8OS*, identify carrier alleles at *RFC1, CSTB and FXN*, and identify a CGG repeat expansion with associated hypermethylation in the 5’ UTR of *PCMTD2* in an incompletely diagnosed individual that shows intergenerational expansion.^31^ TRoLR is freely available at https://github.com/millerlaboratory/TRoLR.

**Figure 1:**
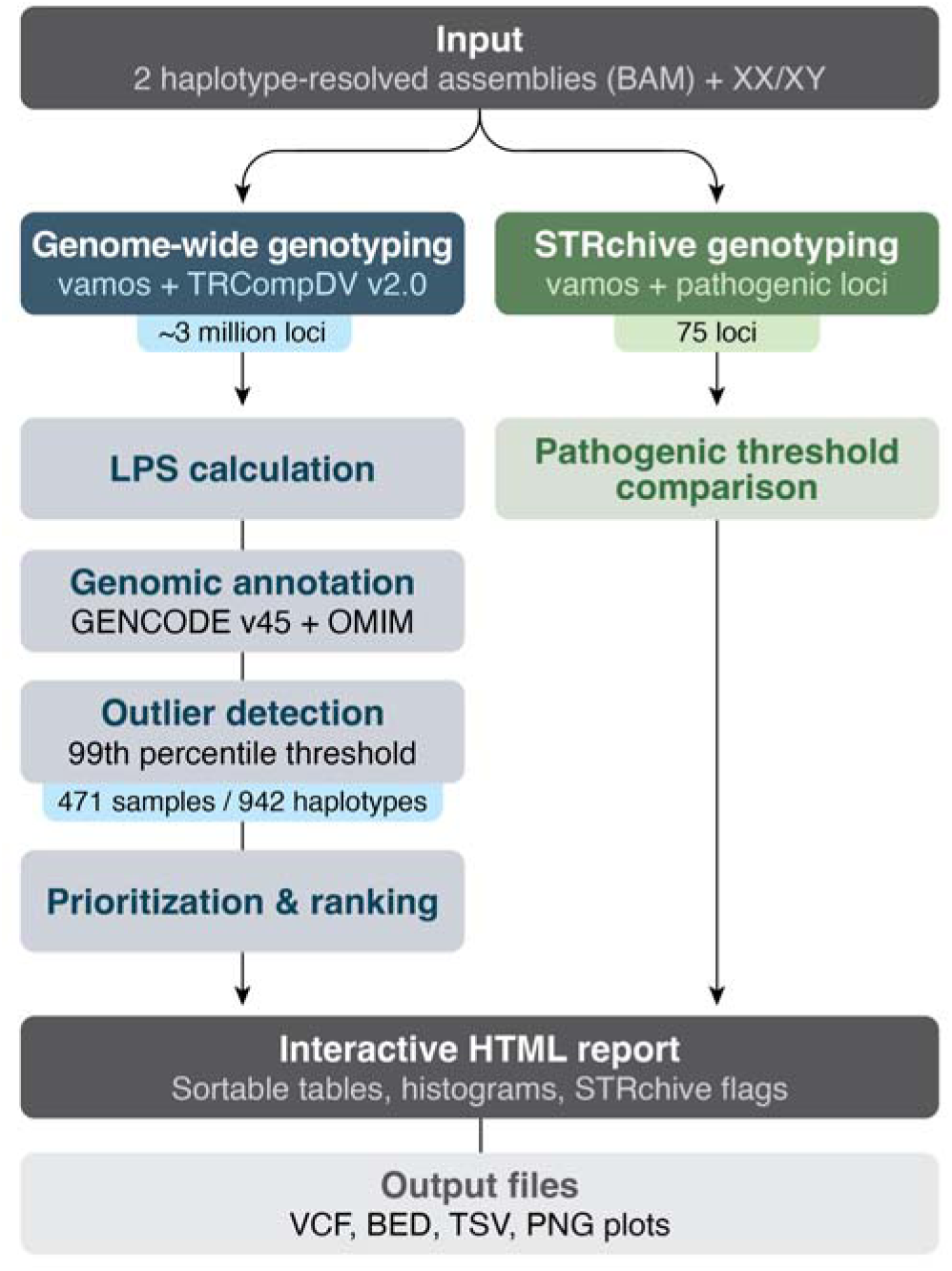
TRoLR pipeline overview. TRoLR takes two haplotype-resolved assemblies (BAM format) aligned to GRCh38/hg38 as input, along with sample karyotype specification (XX or XY). The pipeline genotypes tandem repeats at each haplotype using vamos against both a genome-wide catalog (**Data File 1**) and a curated set of known pathogenic loci from STRchive (**Data File 2**). The longest pure repeat segment (LPS) is calculated for each allele and compared to population reference distributions derived from 942 haplotypes (471 individuals). Outliers exceeding the 99th percentile are annotated with genomic context from GENCODE and compiled into an interactive HTML report. The control database will increase over time as additional samples are sequenced.

## METHODS

### Study cohort

Our reference set includes 242 individuals from the 1KGP-LRSC sequenced using the ONT R10.4.1 pore and 229 individuals from the HPRC Release 2 **(Table S1)**.^12,26^ Samples were selected to represent diverse global populations and included individuals of African, American, East Asian, European, and South Asian ancestry. All 1KGP-LRSC samples had high-coverage sequencing data (>25×) with read N50 exceeding 40 kbp.

We analyzed 47 probands from the UDN who remained undiagnosed or incompletely diagnosed after comprehensive genetic testing that included exome or genome sequencing **(Table S2)**. Informed consent was obtained for all UDN participants under protocols approved by the National Institutes of Health Institutional Review Board (IRB#15HG0130). Long-read ONT sequencing for the UDN samples was performed on blood or fibroblast-derived DNA on a PromethION 24 prepared using the ligation library kit (SQK-LSK114) with the R10.4.1 pore and basecalled using Dorado (ONT).

### Haplotype-aware *de novo* assemblies

Haplotype-resolved *de novo* assemblies were generated for each of 242 1KGP-LRSC samples and the 47 UDN samples using hifiasm (v0.19.5) in ONT mode (--ont) with default parameters.^32^ This produced two haplotype-specific assemblies per sample, yielding 484 total assemblies. 458 HPRC release 2 assemblies, representing 229 individuals, were downloaded from the public repository (https://data.humanpangenome.org/assemblies). Each haplotype assembly was aligned to the GRCh38/hg38 reference genome (https://hgdownload.soe.ucsc.edu/goldenPath/hg38/bigZips/analysisSet/) using minimap2 (v2.24) with the following parameters for assembly-to-reference alignment: `- ax asm20 --cs -t 40`. The resulting alignments were sorted and indexed using samtools (v1.22)^33^. Assembly quality was assessed using quast (v5.2.0).^34^ Assembly quality metrics, including contig N50 and total assembly length, are available **(Table S3)**.

### Tandem repeat genotyping

Tandem repeat genotyping was performed on the 942 aligned assemblies using vamos (v3.0.6) with the `–contig` flag, which is specifically designed for genotyping tandem repeats from aligned genome assemblies rather than read alignments.^11^ We selected TRCompDB v2.0 efficient motif catalog (vamosExpanded_v3.0_GRCh38_effMotifs-0.1.tsv, accessed February 16^th^ 2026) because it provides optimized motif definitions derived from 416 high-quality long-read assemblies that span diverse populations and has overlap with samples genotyped for the underlying TRoLR control cohort.^35^ The catalog was filtered to 3,062,349 loci by excluding homopolymer loci (motif length = 1 bp) and loci overlapping STRchive entries, of which 2,660,635 are STRs and 401,714 are VNTRs with a maximum motif size of 668bp **(Data File 1)**. STRchive loci were genotyped separately using STRchive-specific motif definitions to enable comparison against established pathogenic thresholds. Because Vamos takes in a base-1 locus file unless specified with the `–Z` flag the STRchive loci were genotyped with the `–Z` flag (because they are 0-based), whereas the genome-wide catalog was genotyped with default settings. Both commands include the `–S` and `-E` flags to preserve the native string and the decomposed motif string to retain both the native allele sequence and the decomposed motif string in the output for downstream analysis. Vamos decomposes each allele sequence into motif units by matching segments to the most similar motif in the catalog, an approach that accommodates sequencing errors in more error prone long reads that might otherwise fragment the detected repeat structure. Conversely, this may report non-native sequence including pathogenic-associated motifs that more closely resemble the sequence detected introducing false-positives.

### Longest pure repeat segment calculation

The longest pure repeat segment (LPS) was calculated for each allele at each locus by analyzing the numeric motif-decomposed allele string output by vamos (vamos_lps.py). This is generating using the decomposed allele sequence based on the given motifs for each locus. If the actual sequence is not in the efficient set, it will be corrected to the nearest motif. For a given allele, the LPS metric captures both the repeat length and the presence of interruptions, which can have functional and clinical significance for repeat expansion disorders.

Specifically, for each decomposed allele sequence, we identified and recorded the longest contiguous run of identical motif units. The motif identity associated with the LPS was also recorded, as some loci contain multiple motif types. For example, a CAG repeat array with a CAA interruption would have its LPS calculated as the longest uninterrupted CAG tract rather than the total repeat length. This approach follows the methodology established by the *All of Us* Research Program for characterizing tandem repeat variation.^23,24^ For the control catalog, the LPS is calculated from each haplotype resolved VCF and merged into a single diploid VCF for each sample (lps_per_sample_filter.py). At this step, the longest allele for XY samples is selected in the case where there are two alleles at X-linked loci. Diploid sample VCFs are then combined into a merged VCF for the population (combine_sample_vcf.py).

### Genomic annotation

Loci were annotated from a modified GENCODEv45 file filtered to retain only the canonical transcript isoform for each gene. Annotation was performed using bedtools intersect (v2.31.0) to classify each repeat locus by its genomic context: exonic, intronic, UTR, or intergenic. Loci were only considered exonic if at least 70% of the region overlapped with an exon. This threshold was selected to exclude loci that marginally overlap exon boundaries while retaining loci that are predominantly exonic. For loci spanning multiple annotation categories, priority was assigned in the following order: exon > 5’ UTR > 3’ UTR > intron. Genes associated with a Mendelian disease (as defined by OMIM), not limited to TR-associated disorders, are also annotated with the associated OMIM catalog number and the associated condition and phenotypes.

### Population reference catalog generation

To establish population-level reference distributions for outlier detection, we calculated summary statistics for the LPS at each locus across all 942 haplotypes from the 471 control samples. For each locus, we computed 1) the mean LPS (the arithmetic mean of LPS values across all haplotypes); 2) standard deviation (the standard deviation of LPS values); 3) median LPS (the median LPS value); 4) 99th percentile LPS (the LPS value at the 99th percentile of the distribution); and 5) the range (the 99th percentile LPS minus the minimum LPS). These statistics were calculated separately for autosomal loci and sex chromosome loci to account for ploidy differences. For X chromosome loci, statistics were calculated using all XX samples (two haplotypes each) and the single X chromosome haplotype from XY samples. Y chromosome loci were analyzed using only the XY samples. The resulting population reference catalog contains LPS distributions for over 3 million tandem repeat loci genome-wide and serves as the basis for outlier identification in query samples.

### STRchive pathogenic locus annotation

We performed targeted genotyping at known pathogenic tandem repeat loci catalogued in STRchive (v1.0)**(Data File 2)**.^6^ STRchive is a curated database of disease-associated tandem repeat loci that includes information on pathogenic repeat thresholds, associated phenotypes, inheritance patterns, and molecular mechanisms. We modified the available general STRchive catalog to include additional benign, pathogenic, and interrupting motifs based on compiled information for each locus.^6,36–50^ The LPS summary statistics for each STRchive locus are included in the overall catalog.

Each STRchive VCF is passed through the pathogenic_detector.py script, which uses the total motif count for each allele instead of the LPS against established pathogenic and premutation thresholds because established clinical pathogenic thresholds are typically defined in terms of total repeat count rather than longest pure segment length. Loci exceeding these clinical thresholds are flagged for priority review in the final report. Ten loci were excluded from STRchive reporting due to limitations in interpretability, but genotype data for these loci are available in the VCF files output by the pipeline. Some excluded loci are associated with repeat contractions rather than expansions (*COMP*, *VWA1*, *MIR7-2*), others involve methylation-dependent mechanisms not captured by repeat length alone (*XYLT1*), several have limited or conflicting evidence supporting the repeat expansion as causal (*NIPA1, DMD, POLG, ZIC3*), and two have noted difficulty genotyping which lead to low confidence results (*MUC1*, *FOXL2*).^6,51,52^

### TRoLR pipeline implementation

TRoLR is implemented as a Bash pipeline with supporting Python and R scripts. The pipeline is available at https://github.com/millerlaboratory/TRoLR under the MIT license. TRoLR requires two haplotype-resolved assembly files (BAM format) aligned to GRCh38/hg38 and sample karyotype specification (XX or XY) **(Figure 1)**. The pipeline is designed for haplotype-aware assemblies and is not optimized for samples with aneuploidy or for direct analysis of read alignments.

The TRoLR pipeline first performs genotyping of defined repeats in which each haplotype assembly is processed separately using vamos with the --contig option against both the genome-wide catalog and the STRchive catalog. This generates VCF files containing decomposed repeat alleles for each haplotype. An LPS calculation is then performed where the decomposed allele sequences from the genome-wide VCF files are parsed to calculate the LPS at each locus for each haplotype. Results are output as a BED file with columns for chromosome, start position, end position, LPS length, motif identity, and haplotype indicator **(Table 1)**. Genotypes at known pathogenic loci are then compared to established clinical thresholds from STRchive and flagged if they exceed pathogenic or premutation ranges. The LPS BED file is then intersected with the GENCODE annotation file to add genomic context (exon, intron, UTR) and gene information to each locus. Per locus outliers are identified by comparing the sample’s LPS to the population reference statistics. An outlier is defined as any LPS value exceeding the 99th percentile LPS length established in the control population.

**Table 1.** Output files generated by TRoLR in a sample-specific directory.

| File | Description |
| --- | --- |
| `<sample>_hp1.vcf.gz` | Vamos VCF for haplotype 1 (genome-wide catalog) |
| `<sample>_hp2.vcf.gz` | Vamos VCF for haplotype 2 (genome-wide catalog) |
| `<sample>_hp1.strchive.vcf` | Vamos VCF for haplotype 1 (STRchive loci) |
| `<sample>_hp2.strchive.vcf` | Vamos VCF for haplotype 2 (STRchive loci) |
| `<sample>_lps.bed` | LPS values for all loci, both haplotypes |
| `<sample>_lps_annotated.bed` | LPS values with genomic annotations |
| `<sample>_lps_annotated_outliers.bed` | Loci exceeding 99th percentile threshold |
| `<sample>_pathogenic_results.tsv` | Genotypes at STRchive pathogenic loci with comparison to established thresholds |
| `<sample>_outlier_report.html` | Interactive HTML report with sortable tables and embedded distribution plots |
| `plots/*.png` | Distribution plots for outlier loci |

This threshold was selected based on analysis of known pathogenic repeat expansions, which typically fall well above the 99th percentile in unaffected populations. TRoLR’s outlier detection is designed as a screening and prioritization framework rather than a formal statistical test, and as such does not apply genome-wide multiple testing correction. At a 99th percentile threshold, approximately 1% of genotyped loci are expected to be flagged as outliers in any given sample. To reduce this candidate set to a manageable number for review, TRoLR applies a layered prioritization strategy. Outliers are first stratified by genomic context, with exonic and UTR loci prioritized over intronic and intergenic loci. Within each category, candidates are ranked by expansion magnitude (observed LPS minus 99th percentile). Intronic outliers are further filtered to display only those exceeding 100 repeat units, and loci at known pathogenic sites catalogued in STRchive are evaluated against established clinical thresholds independently of the genome-wide analysis. This prioritization scheme surfaces the most extreme expansions for review.

TRoLR also generates an interactive HTML report to facilitate review and prioritization of outlier expansions **(Figure S1)**. The report includes a summary section which has an overview of total outliers identified, broken down by genomic context (exonic, UTR, intronic, intergenic). Next, the STRchive results section displays results for known pathogenic loci, including comparison to established thresholds. Loci exceeding pathogenic or premutation thresholds are highlighted. Following this, outlier expansions in coding regions, prioritized by expansion magnitude, followed by outlier expansions in 5’ and 3’ UTRs, then intronic outlier expansions, filtered to show the most significant candidates. For each reported outlier, an embedded histogram displays the population LPS distribution at that locus with the sample’s value indicated, enabling visual assessment of outlier magnitude. Given the large number of intronic repeats, the intron section displays only outliers exceeding 100 repeat units. Non-prioritized loci which include smaller intronic repeats and unannotated loci with motifs over 2 bp are browsable in a separate table, as are loci with 2 bp motifs in another table. The report uses sortable and searchable tables implemented in JavaScript to enable rapid filtering by gene name, motif type, or expansion size.

For each prioritized outlier locus, TRoLR generates a histogram plot showing the distribution of LPS values across the reference population **(Figure S1)**. The query sample’s LPS value is indicated by a vertical line, allowing visual assessment of the outlier’s position relative to the population distribution. Plots are generated using R (v4.4.2) with ggplot2 (v3.4.0) and are embedded in the HTML report and saved as individual PNG files.

### gnomad-TR data

STR genotype data from gnomAD-TR was downloaded on March 17^th^ 2026^53^.

### Tandem Repeat Read-Level Genotyping

Reads from the UDN trio were genotyped at the *PCMTD2* expansion using straglr (v1.5).^20^ Results were plotted using the R package ggbreak.^54^

### Software environment and reproducibility

The TRoLR pipeline dependencies are specified in a conda environment file (environment.yaml) included in the repository. Key software versions include: samtools v1.22, vamos v3.0.6, bedtools v2.31.1, Python v3.8+, R v4.0+. The pipeline was developed and tested on Ubuntu 22.04 LTS. Runtime for the TRoLR pipeline itself (excluding assembly generation and alignment) is approximately 5-10 minutes per sample.

## RESULTS

### Tandem repeat landscape in the 1000 Genomes Project cohort

A modified TRcompDB v2.0 catalog was curated to exclude homopolymers and STRchive loci (which are genotyped separately) for analyzing the LPS landscape across 471 publicly available samples (942 haplotype-resolved assemblies) from the 1KGP-LRSC and the HPRC. We obtained genotyping data at 3,062,232 of the 3,062,349 loci in the catalog (99.9%). While only 5,076 autosomal loci captured genotype information for all expected 942 alleles, the median genotyped allele fraction is 99.3% with 96.2% of autosomal loci containing at least one haplotype for each sample **(Data File 3)**. For the STRchive loci, no autosomal locus captured all haplotypes, but all but one had at least 1 allele per-sample with a median fraction of 99.1% of alleles genotyped. For both catalogs, the median fraction of alleles genotyped at X-linked loci was 99.1% **(Data File 4)**. The distribution of tandem repeat loci across genomic regions reflected the underlying genome architecture: 64.5% of loci were in intergenic regions, 32.8% in introns, 1.25% in exons, 1.10% in 3’ UTRs, and 0.36% in 5’ UTRs **(Figure 2A)**.

**Figure 2:**
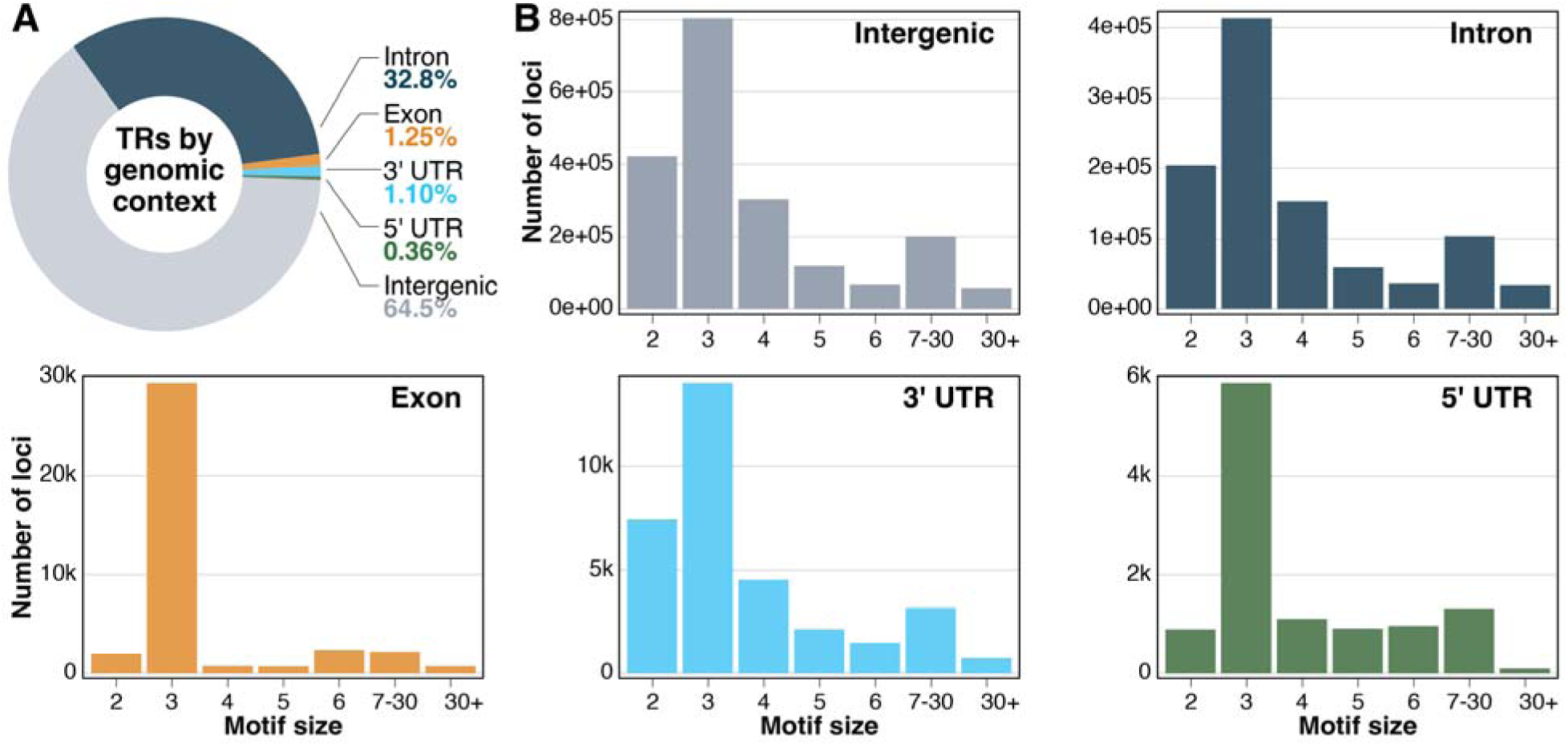
Genomic distribution of tandem repeat loci in the TRCompDB v2.0 catalog. **(A)** Proportion of loci annotated as exonic (1.25%), 5’ UTR (0.36%), 3’ UTR (1.10%), intronic (32.8%), or intergenic (64.5%). **(B)** Distribution of motif sizes stratified by genomic annotation context.

Across all contexts, trinucleotide repeats represent the most common motif size representing 41% of the catalog with dinucleotide repeats the second most common, making up 21% of the catalog **(Figure 2B)**. The curated catalog incorporates the TRExplorer catalog, which is enriched for short, pure repetitive sequences. This composition likely contributes to the high prevalence of trinucleotide repeats in the catalog, particularly among exonic loci.^29,35^ Among trinucleotide repeats, CAG/CTG (the motif class associated with polyglutamine repeats) was the most abundant motif, representing 25% of exonic trinucleotide loci which is clinically important because CAG expansions underlie polyglutamine disorders.

### Population-level and population-specific LPS variation

To establish reference distributions for outlier detection, we calculated summary statistics for the LPS at each locus across all haplotypes **(Data Files 5 and 6)**. We observed substantial variation in repeat length distributions, with the median LPS ranging from 1 to 1165 motif units depending on repeat class and genomic context **(Figure 3A)**. The coefficient of variation (CV) for LPS ranged from 0 to 23.6 across loci, with 74.3% of loci having a CV of 0 and 25.7% being polymorphic. This demonstrates that most TR loci in the genome have little to no variability across the control set, though noncoding and intronic repeat loci had higher variance on average compared to exonic repeats (Two sample Wilcoxon rank-sum test *p*=7.44e-85) where the coefficient of variation was greater than 0 **(Figure 3C)**.

**Figure 3.**
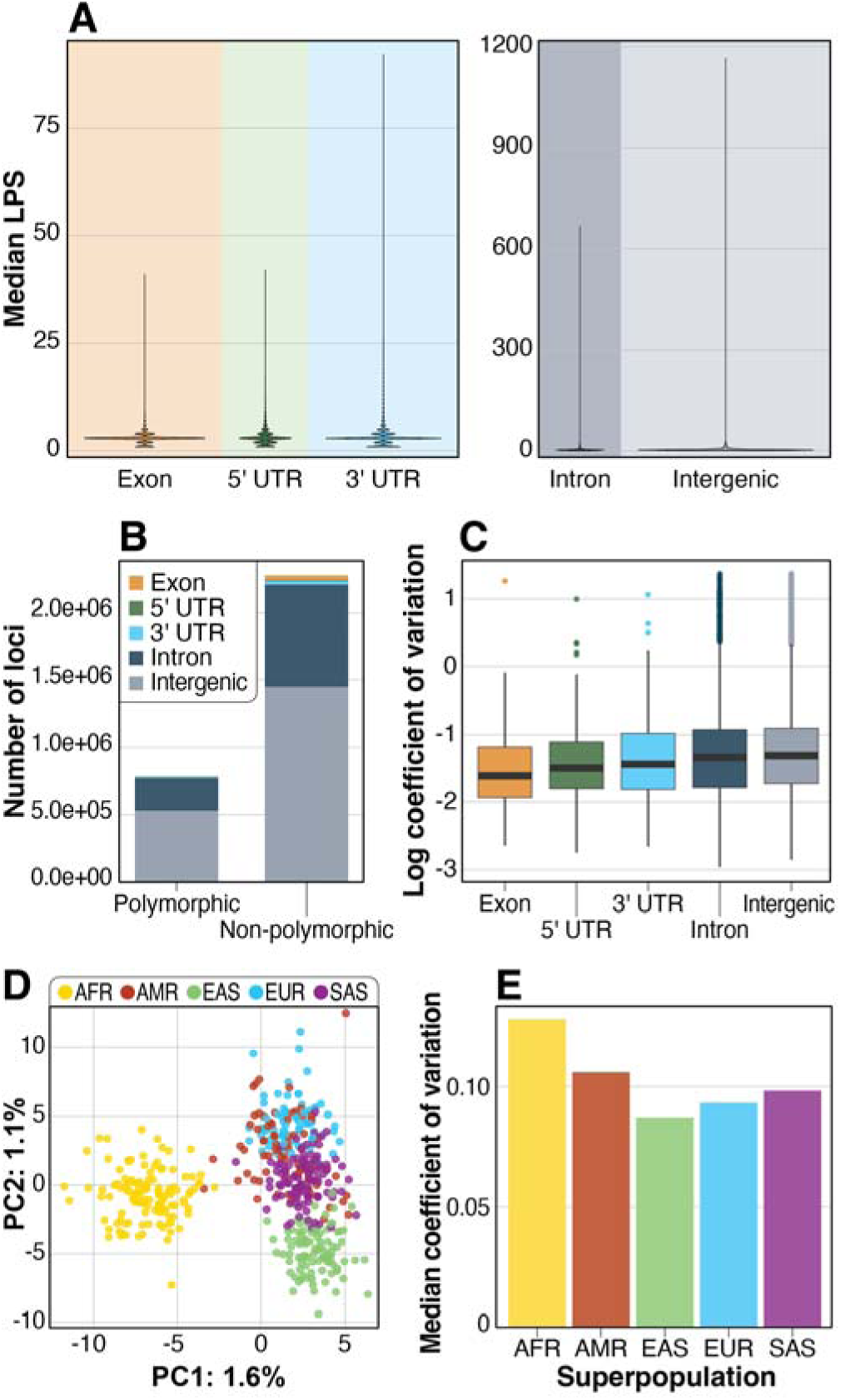
Population-level LPS variability across 942 long-read genome assemblies. **(A)** Violin plots representing the distribution of median LPS values stratified by genomic annotation. **(B)** Stacked bar plot showing the number of polymorphic (coefficient of variation [CV] > 0) and non-polymorphic (CV = 0) loci in each annotation category. **(C)** Box plot of log-scaled CV for polymorphic loci, stratified by genomic annotation. **(D)** Principal component analysis of tandem repeat variation across superpopulations, performed using 2,128 loci with SD > 0 and complete genotype information across all 942 haplotypes. **(E)** Median CV for each superpopulation across 214,677 loci showing statistically significant population stratification (One-way Kruskal-Wallis test, Bonferroni-corrected p < 0.05).

Using principal component analysis of 2,128 loci with non-zero standard deviation and complete genotype information, we confirmed that polymorphic tandem repeats can segregate samples by superpopulation **(Figure 3D)**^55^. For the majority of loci (93%), LPS distributions did not differ significantly across superpopulations (One-way Kruskal-Wallis test, Bonferroni-corrected *p* > 0.05). However, 214,677 loci (7%) showed statistically significant population stratification (Bonferroni-corrected *p* ≤ 0.05), consistent with the greater genetic diversity observed in African ancestry populations across other variant classes **(Data File 7, Figure 3E)**^56^.

### Tandem repeat outlier detection

To establish an outlier detection threshold, we evaluated the sensitivity of percentile-based cutoffs for known pathogenic repeat expansions catalogued in STRchive. At the 99th percentile threshold, most control cohort samples are below the established pathogenic thresholds **(Figure S2)**. This does not hold true at several complex loci where specific motifs are associated with pathogenicity and benign motifs can expand beyond the known threshold, such as *DAB1.* We therefore selected the 99th percentile as a threshold that balances sensitivity for clinically relevant expansions against a manageable number of candidates for review, while acknowledging that this threshold is empirically calibrated and may warrant adjustment as larger reference populations and additional validated pathogenic loci become available.

Using this threshold, TRoLR identified a median of 2,144 outlier loci per cohort sample (approximately 0.07% of loci genotyped, range: 1,266–8,073) across all genomic contexts. The number of outliers varied by genomic region: individuals had a median of 7 exonic outliers, 9 5’ UTR outliers, 20 3’ UTR outliers and 667 intronic outliers. When stratified by superpopulation, individuals of African ancestry had the highest median number of outliers per sample **(Figure 4A)**, a pattern consistent with the greater genetic diversity observed in African ancestry populations and with the higher per-locus LPS variance we observed in this group.

**Figure 4.**
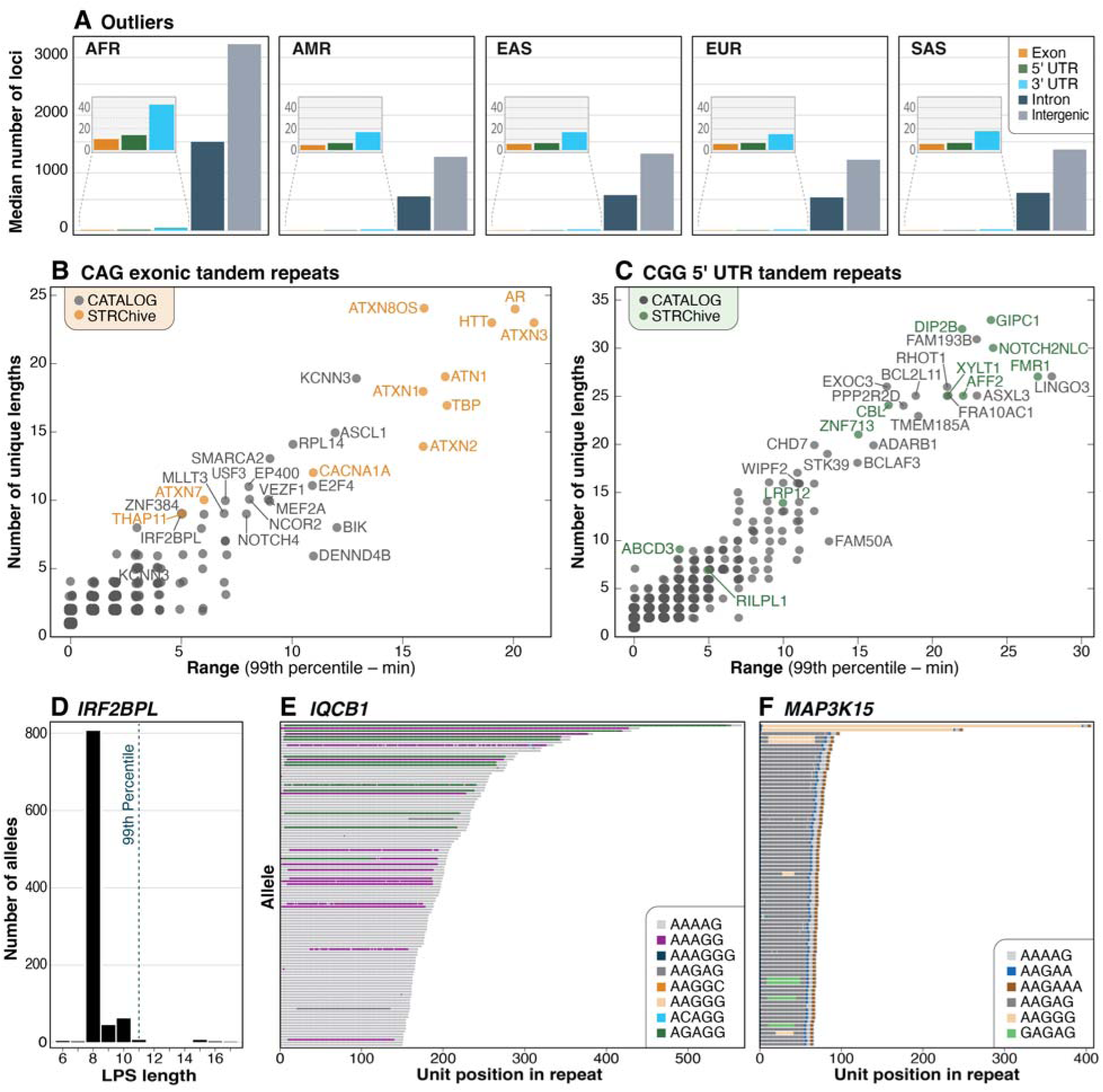
LPS length outliers detected within the 942-haplotype cohort. **(A)** Median number of loci detected per sample per annotation feature stratified by superpopulation. **(B,C)** Dot plots of the range (99^th^ percentile – min) vs the number of unique LPS lengths for **(B)** CAG exonic loci and **(C)** CGG 5’ UTR loci. **(D)** LPS distribution for the CAG tandem repeat in the exon of *IRF2BPL* with the 99^th^ percentile value identified by the red line. **(E)** Waterfall plot of long alleles (≥150 units) in the intronic pentanucleotide repeat in *IQCB1*. **(F)** Waterfall plot of long alleles (≥66 units) in the intronic pentanucleotide repeat in *MAP3K15*.

To assess the variability among exonic CAG and 5’ UTR CGG tandem repeats within our control cohort, we examined the calculated range (99^th^ percentile minus the minimum LPS values) versus the number of unique LPS lengths and identified outliers at both previously known pathogenic loci and novel loci not currently associated with a disease, similar to recent findings in the *All of Us* cohort **(Figure 4B,4C)**.^16^ Importantly, this analysis identifies loci that may have the propensity to expand and should be prioritized in studies looking for novel disease-associated expansions. As an example, *EP400, FAM193B,* and *BCLAF3* are all outliers, and pathogenic alleles have recently been proposed for all three loci but they are not yet listed in STRchive.^23,57,58^

We then examined the overlap between outliers within our control cohort and pathogenic loci from STRchive to evaluate the sensitivity. Of the 75 STRchive loci genotyped across all samples, at least one individual exceeded the 99th percentile threshold at 58 loci (∼77%) **(Table S4)**. This included the detection of known pathogenic alleles in our samples. For example, similar to a prior report, we identified an expansion of the CAG repeat within *HTT* in HG02470 that is 42 units long—three units longer than the defined pathogenic threshold of 39 (**Figure S3)**.^59^ Conversely, we detected additional expansions associated with autosomal dominant conditions that to our knowledge have not been previously reported, including an expansion in *DIP2B* (NA20752) and three individuals with expansions in *ATXN8OS* (HG02055, HG02258, HG02514), which is known to be a difficult locus to accurately characterize (**Figure S3**).^16^ As no phenotypic data is available for individuals from the 1KGP cohort we are unable to determine the impact, if any, of these expansions in these individuals.^6^

We identified one expanded allele each in the *CSTB*, *AR,* and *TYMS* loci, which are associated with autosomal recessive or X-linked recessive conditions **(Figure S4).** Identification of expanded alleles in *CSTB* and *AR* was consistent with expected population carrier frequencies. Based on gnomAD-TR estimates, the expected number of carriers in our cohort was 0.35 for *CSTB* and 0.14 for *AR* (considering XX individuals only for *AR*; comparable population frequencies are not available for *TYMS*)^53^. Exact binomial testing showed that observing at least one carrier was not significantly greater than expected (*CSTB* p=0.293; *AR* p=0.129), indicating these findings are consistent with population carrier frequencies. We observed 6 alleles in 6 different individuals over the pathogenic minimum at *RFC1* and with the pathogenic AAGGG motif. Across the control cohort we detected expanded alleles at several loci associated with autosomal dominant conditions, including *ATXN10*, *BEAN1, DAB1, STARD7, MARCHF6, TNRC6A* and *RAI1*, but with a benign motif. Similar expanded alleles from long-read data have been previously reported in individuals from this cohort **(Figure S5)**.^12,19^

Finally, as an example of TRoLR’s ability to detect very large expansions with associated epigenetic changes in the control cohort, we identified one individual (HG00653) who carried a 5′UTR *GIPC1* repeat expansion of 571 GGC units, approximately fivefold larger than the reported pathogenic range (73–164 repeats), with associated hypermethylation **(Figure S6)**.^60^ Expansions at this locus cause oculopharyngodistal myopathy type 2 (OPDM2, MIM: 618940) and are predominantly observed in individuals of East Asian ancestry, consistent with the reported ancestry of this individual.^61,62^ These expansions occur within an upstream open reading frame and produce a polyglycine-containing protein that accumulates in p62-positive inclusions and drives neurodegeneration in model systems.^63^ Individuals with expansions over 500 units long have been reported to be asymptomatic.^62^ The expansion and hypermethylation observed here could reflect somatic expansion during cell line passaging rather than the individual’s germline genotype.

### Exonic repeat outliers

Exonic repeat expansions are of clinical interest because of their potential to directly alter mRNA and protein sequence. Across all samples, we identified 2,309 exonic trinucleotide repeat outliers spanning 384 unique genes at loci not previously associated with TRE disorders. GGC repeats (encoding polyglycine and polyalanine tracts, among others, depending on reading frame) and CAG repeats (encoding polyglutamine tracts) were the most common, representing 37% and 27% of exonic outliers, respectively **(Table 2, Table S5).**

**Table 2.** Top exonic CAG repeat outliers by expansion magnitude.

| Gene | Sample | Genomic location (hg38) | Reference Mean LPS | SD | Sample LPS | 99<sup>th</sup> percentile LPS | Sample LPS minus 99th percentile |
| --- | --- | --- | --- | --- | --- | --- | --- |
| ***PRSS21*** | HG02380 | chr16:2817283-2817309 | 9.0 | 0.45 | 21 | 9 | 12 |
| ***KCNN3*** | HG03788 | chr1:154869724-154869766 | 19.1 | 2.19 | 31 | 23 | 8 |
| ***POU6F2*** | HG01981 | chr7:39339668-39339719 | 10.0 | 0.27 | 18 | 10 | 8 |
| ***VEZF1*** | HG02818 | chr17:57979244-57979283 | 14.0 | 0.61 | 22 | 15 | 7 |
| ***MN1*** | HG02583 | chr22:27798893-27798974 | 9.1 | 0.63 | 17 | 10 | 7 |
| ***BIK*** | HG00625 | chr22:43129229-43129274 | 17.9 | 0.84 | 25 | 18 | 7 |
| ***EP400*** | HG01686 | chr12:132062524-132062607 | 28.7 | 0.92 | 37 | 31 | 6 |
| ***IRF2BPL*** | HG02555 | chr14:77027411-77027535 | 8.3 | 0.93 | 17 | 11 | 6 |

To prioritize candidate loci, we ranked exonic CAG/CTG-repeat outliers based on the magnitude of difference from the 99th percentile threshold. The largest outliers were observed in *PRSS21, KCNN3, POU6F2,* and *IRF2BPL.* Although these genes have not previously been associated with repeat expansion disorders, several have biological roles consistent with neurological disease mechanisms^64^.

Nine individuals in the control cohort carried CAG repeat alleles in *IRF2BPL* exceeding the 99th percentile, consistent with a normal LPS distribution and suggesting that this locus contains a polymorphic polyglutamine tract rather than a rare pathogenic expansion **(Figure 4D)**. *IRF2BPL* encodes a transcriptional regulator, and heterozygous loss-of-function variants cause an autosomal dominant neurodevelopmental disorder characterized by intellectual disability, seizures, and motor delay (MIM: 618088).^64^ While a polymorphic CAG repeat in *IRF2BPL* has been documented, expansions have not previously been implicated in disease.^64,65^ Similarly, we observed eight individuals with outlier-length CAG repeats in *KCNN3*, making this one of the most polymorphic exonic CAG-repeat loci in the cohort **(Figure 4B)**. Variable-length CAG repeats in *KCNN3* have been investigated previously in association with schizophrenia, although results have been inconsistent across studies.^66–68^

### 5’ UTR repeat outliers

The convergence of multiple disease mechanisms at 5’ UTR CGG loci combined with recent discoveries of novel pathogenic expansions at loci such as *FAM193B* suggests that additional disease-associated 5’ UTR CGG expansions likely remain to be identified. One common mechanism is that CGG expansions above a critical threshold trigger DNA hypermethylation of the adjacent CpG island and promoter, leading to transcriptional silencing and loss of gene expression. This methylation-dependent silencing mechanism has been demonstrated at multiple 5’ UTR CGG loci, including *FMR1, XYLT1, and AFF2*^69,70^. These repeats can also be pathogenic through repeat-associated non-AUG (RAN) translation, which causes toxic polyglycine-containing proteins from upstream open reading frames, and through RNA-mediated toxicity, as observed in *FMR1* premutation carriers.^43,63,70^

We identified 2,161 CGG repeat outliers in 5’ UTRs, including at *FRA10AC1, TBC1D7, and FAM193B* **(Table 2**, **Table 3, Figure S7, Table S6)**. Among these, one individual (HG02300) carried a CGG expansion of 589 repeat units at *FRA10AC1*, a gene located at the folate-sensitive fragile site FRA10A and associated with an autosomal recessive neurodevelopmental disorder (MIM: 620113). Expansions exceeding approximately 200 CGG units at this locus have been shown to trigger CpG island hypermethylation and transcriptional silencing, consistent with the methylation-dependent mechanism described at *FMR1*^71^. The expansion in HG02300 was associated with hypermethylation of the surrounding region, corroborating these prior reports.^71^

**Table 3.** Top 5’ UTR CGG repeat outliers by expansion magnitude. Values represent total repeat count. Loci are ranked by the difference between the observed value and the 99th percentile in the reference cohort.

| Gene | Sample | Genomic location (hg38) | Reference Mean LPS | SD | Sample LPS | 99<sup>th</sup> Percentile LPS | Sample LPS minus 99th percentile |
| --- | --- | --- | --- | --- | --- | --- | --- |
| ***FRA10AC1*** | HG02300 | chr10:93702491-93702548 | 8.8 | 19.3 | 589 | 26 | 563 |
| ***TBC1D7*** | HG00115 | chr6:13328502-13328533 | 4.4 | 10.0 | 311 | 6 | 305 |
| ***PCMTD2*** | HG00160 | chr20:64255724-64255749 | 10.3 | 7.4 | 226 | 16 | 210 |
| ***CSNK1E*** | HG04157 | chr22:38317348-38317375 | 10.0 | 6.2 | 194 | 16 | 178 |
| ***FAM193B*** | HG03743 | chr5:177554490-177554531 | 17.1 | 6.0 | 147 | 33 | 114 |

At *FAM193B*, one individual (HG03743) carried an outlier CGG expansion of 147 repeat units, compared to a population 99th percentile of 33 units. Two recent studies have independently proposed that large CGG expansions in the 5’ UTR of *FAM193B* cause a form of oculopharyngodistal myopathy (OPMD), with reported pathogenic alleles over 200 units.^23,57^ The allele observed in our control cohort is substantially shorter than these pathogenic alleles but exceeds the normal range, raising the question of whether it represents a premutation-range allele with potential for intergenerational expansion.

Finally, at *TBC1D7*, one individual (HG00115) carried a CGG expansion of 311 repeat units, well above the population 99th percentile of 6 units. *TBC1D7* encodes a component of the TSC1-TSC2 complex that negatively regulates mTOR signaling. Biallelic loss-of-function variants cause an autosomal recessive megalencephaly syndrome (MIM: 612528), and expansions at this locus were also recently linked to OPMD^72^. Surprisingly, while the expansion in HG00115 is larger than those previously reported (between 83 and 148 units), it remains unmethylated. This is converse to the observed extreme outlier in *GIPC1* (also a cell line) and a similarly sized allele in the study (from a blood sample)^72^. With the continued identification of 5’ UTR CGG OPMD-associated tandem repeats, wider studies are likely needed to establish if there is a shared mechanism across loci and the functional effects of larger expansions.

### Intronic repeat outliers

We focused on pentanucleotide repeats because pathogenic motif-switching at intronic loci, exemplified by *RFC1*, predominantly involves pentanucleotide motifs. This strategy prioritizes loci where motif changes coincide with repeat expansions, similar to the pathogenic motif-switching observed at *RFC1* in cerebellar ataxia, neuropathy, and vestibular areflexia syndrome (CANVAS), as well as recently identified loci such as *SH3RF3* and *ARHGEF3.*^55^ Using this approach, we identified several expanded alleles in intronic pentanucleotide repeats at multiple loci, including *IQCB1* and *MAP3K15* **(Table S7)**.

At *IQCB1*, the reference motif catalogued by vamos is AAAAG, which is a benign motif at *RFC1*. However, inspection of the consensus repeat sequence indicated that the expanding motif in our cohort was AGAGG, which is a motif found in pathogenic alleles *RFC1* associated with CANVAS. Re-genotyping this locus using additional RFC1 pathogenic motifs from STRchive confirmed that the longest alleles were composed primarily of AGAGG, with intermediate-length alleles containing AAAGG and shorter alleles frequently containing AAGAG **(Data File 8, Figure 4E)**. Because pathogenic variants in *IQCB1* cause Senior–Løken syndrome, an autosomal recessive disorder, motif-specific repeat expansions at this locus could plausibly follow a similar inheritance pattern to that observed for RFC1-associated disease.

Similarly, the intronic repeat locus at *MAP3K15* showed evidence of motif switching. The reference motif at this locus is AAGAG; however, consensus sequence inspection revealed that expanded alleles were composed of AAGGG, another pathogenic motif at *RFC1*. Re-genotyping with additional *RFC1*-associated motifs confirmed one XY individual with an AAGGG expansion and two XX individuals with AAGGG expansions exceeding 200 repeat units **(Data File 8, Figure 4F)**. Given that *MAP3K15* resides on the X chromosome, large expansions at this locus may follow an X-linked recessive inheritance pattern if pathogenic.

### Detection of repeat expansions at known pathogenic loci in UDN cases

To assess the clinical utility of TRoLR, we applied the pipeline to two positive control sample from the UDN as well as 45 UDN probands who remained undiagnosed or incompletely diagnosed after standard clinical testing **(Table S8)**. The first positive control case had a previously clinically confirmed intronic pathogenic repeat expansion in *CNBP*. The long-read assembly was concordant with prior clinical testing, characterizing the allele as 215 CAGG repeat units—well above both the established pathogenic threshold of 75 units and the observed 99^th^ percentile of 23 units **(Figure 5A, Figure S8)**. In the second case, TRoLR correctly identified a PCR-confirmed 261 CTG-long expansion in *ATXN8OS* associated with spinocerebellar ataxia type 8 **(Figure 5B)**^73^.

**Figure 5.**
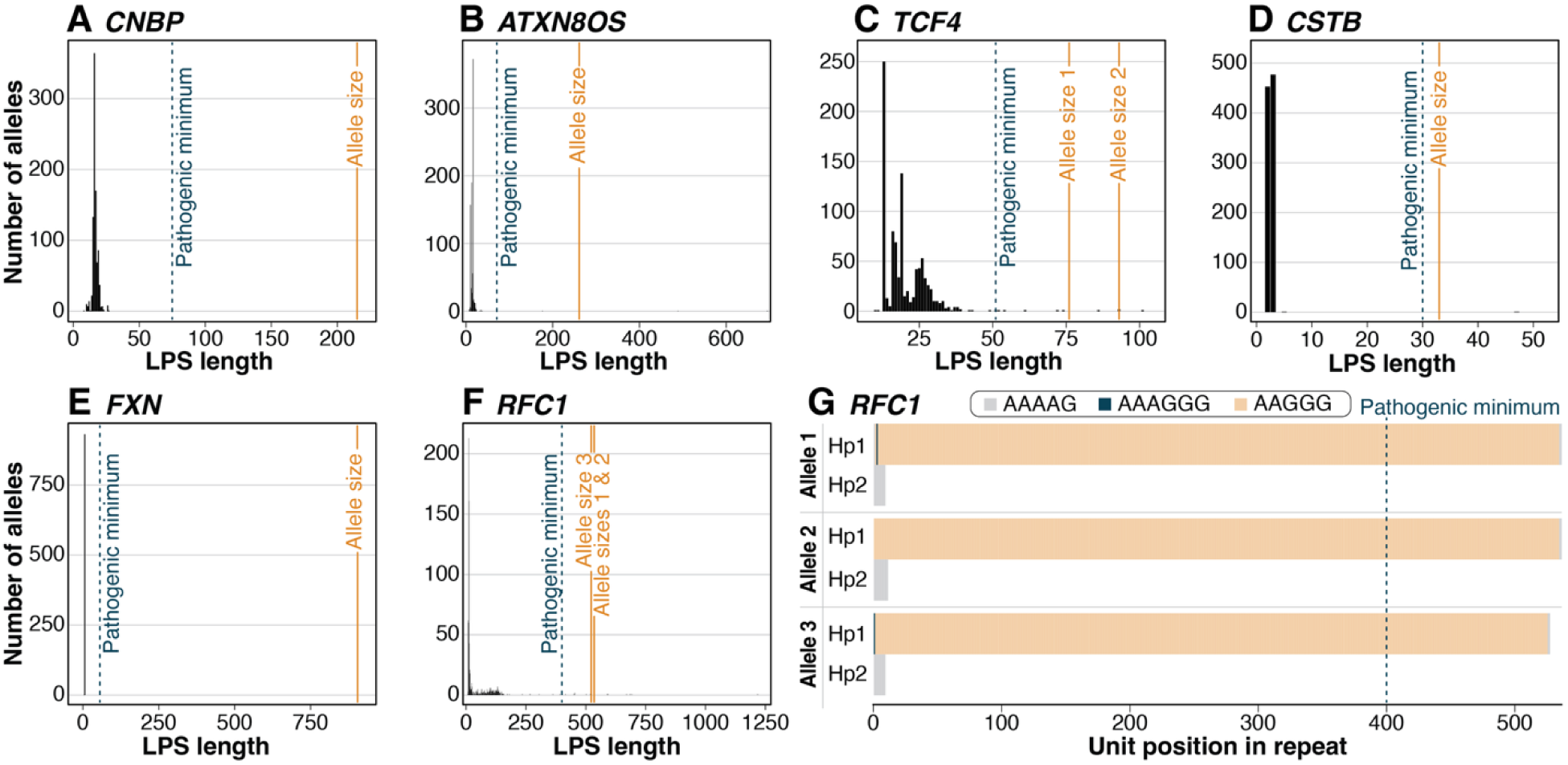
Detection of repeat expansions at known pathogenic loci in UDN cases. **(A–F)** Histograms of LPS distributions from the 942-haplotype reference cohort at five known pathogenic loci: *CNBP*, *ATXN8OS*, *TCF4*, *CSTB*, *FXN*, and *RFC1*. Established pathogenic minimum (dashed line) is indicated for each gene as well as the expanded allele sizes (orange lines) identified in eight samples. **(G)** Modified waterfall plot shows both haplotypes (Hp1, Hp2) for the three individuals with heterozygous AAGGG expansions in *RFC1*. Each horizontal bar represents one haplotype, with segments colored by repeat motif identity.

We then evaluated TRoLR’s ability to detect repeat expansions at loci catalogued in STRchive by analyzing the 45 additional UDN cases sequenced on the ONT platform, none of which had been previously diagnosed with repeat expansion disorders. TRoLR detected repeat expansions at known pathogenic loci in 7 of these individuals. These included a pathogenic-length allele at *TCF4* in three individuals, consistent with the prevalence of this expansion, which also exhibits incomplete penetrance and presents as a late onset, mild phenotype **(Figure 5C).**^6,74^ We also identified alleles of pathogenic length for three autosomal recessive conditions, including one individual with an expanded allele at *CSTB*, which causes progressive myoclonic epilepsy type I **(Figure 5D)**; one individual with an expanded allele at *FXN*, which causes Friedreich’s ataxia **(Figure 5E)**; and three individuals with expanded AAGGG sequences at *RFC1* **(Figure 5F,G)**. Carrier frequencies for *FXN* and *RFC1* are relatively high (approximately 0.4–4%, depending on the population) and detection of these alleles in this cohort is therefore consistent with expected background carrier variation. These findings demonstrate TRoLR’s ability to detect candidate pathogenic expansions at established disease-associated loci in previously unsolved cases.

### Identification of a novel 5’ UTR expansion in an incompletely solved UDN case

In one of the 45 UDN individuals, TRoLR prioritized 21 outlier loci, including exonic and UTR outliers with motifs of at least 3 bp and intronic outliers with at least 100 repeat units **(Figure 6A)**. This list included a large CGG repeat expansion in the promoter/5′UTR region of *PCMTD2*, a gene that is not included in STRchive or associated with a Mendelian condition. The proband carried an allele of 879 repeat units—substantially larger than both the population median (10 repeats) and the 99th percentile (17 repeats) **(Figure 6B).** Examination of this region revealed hypermethylation of the haplotype across the repeat-containing region **(Figure 6C)**. In the control cohort, one individual (HG00160) carried a shorter expansion (∼226 repeats) at this locus that was also hypermethylated, suggesting that repeat-length-dependent methylation occurs at this locus.

**Figure 6.**
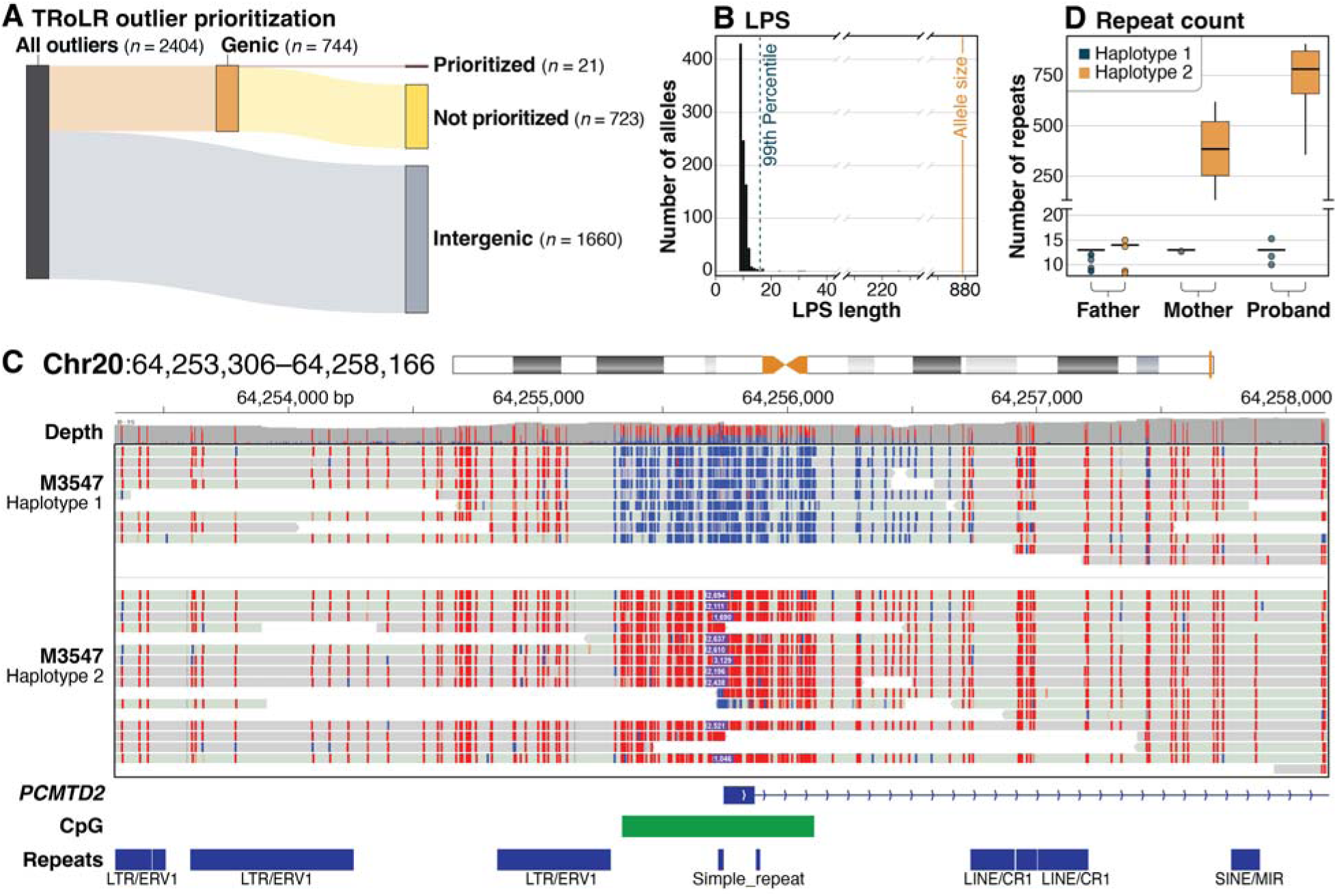
A novel CGG repeat expansion in *PCMTD2* identified in a UDN case. **(A)** Sankey diagram illustrating the TRoLR outlier prioritization workflow for this individual shows the number of outlier loci filtered at each step from genome-wide calls to the prioritized candidates, which included *PCMTD2*. **(B)** Histogram of LPS values at the *PCMTD2* locus across the reference cohort. The dotted line indicates the 99th percentile (17 repeats); the orange line indicates the proband’s expanded allele (∼879 repeats). **(C)** IGV visualization of nanopore reads spanning the *PCMTD2* repeat region in the proband and colored by per-read 5-methylcytosine probability, demonstrating hypermethylation of the expanded allele. **(D)** Box plot of read-level repeat counts at the *PCMTD2* locus for each haplotype for the proband and both parents, showing intergenerational expansion from the maternal allele.

*PCMTD2* encodes a predicted protein-L-isoaspartate O-methyltransferase that may participate in ubiquitination-associated protein turnover. Because CGG-repeat expansions in promoter and 5′UTR regions are known to alter gene expression through local hypermethylation, we considered whether this expansion could contribute to the individual’s phenotype. Sequencing of the unaffected parents revealed that the mother also carried a shorter expanded allele (∼504 repeats), approximately 380 repeat units shorter than observed in the proband **(Figure 6D)**, demonstrating intergenerational repeat instability similar to that observed at other CGG-repeat loci^58,70^. Although the heterozygous expansion in this individual remains a variant of uncertain significance, as *PCMTD2* has not yet been clearly associated with a Mendelian disorder, this finding illustrates the potential of TRoLR to identify candidate pathogenic repeat expansions in previously unsolved cases.

## DISCUSSION

Our analysis of 471 diverse individuals from the 1000 Genomes Project establishes population reference distributions for over 3 million tandem repeat loci and demonstrates the utility of this approach for both research discovery and clinical diagnostics. This work revealed statistically significant differences in LPS distributions at 7% of loci across the five superpopulations in our reference cohort, with individuals of African ancestry exhibiting the greatest median coefficient of variation, consistent with the higher genetic diversity observed in this group. This pattern represents genuine population-level variation rather than technical artifact, and it underscores the importance of ancestry-diverse reference panels for calibrating outlier thresholds. Individuals from populations underrepresented in the reference cohort may be disproportionately flagged as outliers if population-specific allele frequency distributions are not accounted for. While our cohort provides an initial foundation for ancestry-informed outlier detection, larger and more diverse LRS reference datasets—such as those being generated by the *All of Us* Research Program—will be essential for refining population-specific thresholds^23,24^.

By integrating haplotype-aware assembly-based genotyping, population-level outlier detection using the LPS, and clinical prioritization with genomic annotation and STRchive thresholds, TRoLR addresses a critical gap between raw long-read data and actionable variant calls for TREs. A distinguishing feature of TRoLR is the use of the LPS rather than total repeat length for outlier detection, which has important biological and clinical implications. By quantifying the longest uninterrupted tract, TRoLR captures this essential dimension of repeat structure. Our population-level LPS distributions can therefore identify repeat outliers and suggest the loss of stabilizing interruptions.

The use of genome assemblies rather than read alignments offers several advantages for TRE analysis. Assemblies provide a consensus representation of each haplotype, reducing the noise inherent in individual long reads. The increasing maturity of assembly tools—including hifiasm, which now supports ONT data—makes this approach practical for clinical implementation^32^. However, assembly-based analysis sacrifices read-level information, including DNA methylation and somatic mosaicism, that may be clinically important. We therefore recommend that outliers identified by TRoLR be reviewed in read-level alignments to confirm that the assembly is representative and to enable downstream epigenetic analysis.

Our analysis of UDN cases demonstrates TRoLR’s potential clinical value. The pipeline confirmed two known expansions concordant with prior testing, identified carrier-range alleles consistent with expected population carrier frequencies, and detected the novel *PCMTD2* expansion in an incompletely solved case. The 5’ UTR CGG expansion follows a common epigenetic paradigm in known pathogenic loci. Although *PCMTD2* is not yet associated with a Mendelian disorder, the observation of repeat-length-dependent methylation in both the proband and an unrelated individual in the control cohort suggests a consistent epigenetic consequence of expansion at this locus. Together, these findings illustrate TRoLR’s ability to both confirm known pathogenic variants and discover candidate expansions at loci not included in current clinical panels, extending the diagnostic reach of LRS beyond what is achievable with targeted repeat analysis alone.

In addition to confirming known pathogenic expansions, TRoLR identified several other candidate loci warranting further investigation. The intronic pentanucleotide expansions at *IQCB1* and *MAP3K15* within our control cohort, were composed of motifs associated with pathogenicity at *RFC1*, suggesting that the motif-switching mechanism underlying CANVAS may operate at additional genomic loci. It is worth noting that the existence of these alternative alleles required manual inspection of the consensus sequences since they were not part of the efficient motif set. This highlights a limitation in vamos as it cannot discover new motifs that are not already in the catalog. Further k-mer analysis may be required to uncover more alternative motif sequences not currently represented in catalogs. Among exonic outliers, the polymorphic CAG tracts in *IRF2BPL* and *KCNN3* highlight the challenge of distinguishing pathogenic expansions from benign population variation at the tail of normal distributions. The human genome contains over 60 genes with polymorphic exonic CAG repeats, and it would be surprising if all pathogenic polyglutamine disorders have already been identified^75^. Systematic assessment of these loci in large clinical cohorts with phenotypic data will be needed to determine whether any harbor pathogenic expansions. Our observation of outlier alleles at loci not yet listed in STRchive, including *EP400*, *FAM193B*, and *BCLAF3*, where pathogenic expansions have recently been reported by others, demonstrates that TRoLR can identify candidate disease loci independently of existing curated databases^23,57,58^. This genome-wide discovery capability complements the targeted assessment at known pathogenic loci.

Several limitations should be acknowledged. First, although ONT R10.4.1 chemistry represents a substantial improvement in accuracy over earlier generations, systematic sequencing errors persist^76,77^. These errors can affect LPS calculation, potentially leading to underestimation of pure tract length or false inference of interruptions. The use of assembly consensus sequences in our control set mitigates but does not eliminate this issue, and direct benchmarking against orthogonal methods (e.g., Southern blot or long-range PCR) was not performed.

Second, our reference cohort of 471 individuals provides limited statistical power for establishing robust 99th percentile thresholds, particularly at highly polymorphic loci where the upper tail of the distribution may be poorly estimated. The empirically chosen 99th percentile threshold balances sensitivity and specificity for known pathogenic expansions in our validation but may not be optimal for all contexts. TRoLR is designed to accommodate updated reference catalogs from both ONT and PacBio data, which we will add as larger LRS population datasets become available.

Third, assembly-based analysis cannot currently detect expansions that exceed the contiguity of the assembly. While modern assemblies can span most clinically relevant regions, very large expansions (e.g., >10,000 repeat units) may be fragmented across contigs or absent from the assembly entirely^12^. Generating accurate consensus sequences may also fail in complex regions of the genome, including segmental duplications. Reviewing read-level alignments remains necessary for the most extreme expansions.

Finally, the 99th percentile threshold is a prioritization filter rather than a formal significance test, and outlier status at any individual locus should not be interpreted as evidence of pathogenicity without additional clinical and functional assessment. The expected false positive rate—defined here as outlier calls at loci with no clinical relevance—is inherently high by design, as the pipeline intentionally casts a wide net to avoid missing true expansions. The layered prioritization workflow, rather than statistical correction, is the mechanism by which the candidate list is narrowed to a tractable number.

As LRS is increasingly adopted in clinical genetics, standardized and automated analysis pipelines will be essential for ensuring reproducibility and enabling data sharing across institutions. TRoLR provides a foundation for systematic tandem repeat analysis in this context, enabling both confirmation of known pathogenic variants and discovery of novel repeat expansions in individuals with suspected genetic disorders. It is designed to integrate with existing clinical LRS workflows. The pipeline accepts assembly-aligned BAM files as input, produces outputs in familiar formats (VCF, BED, TSV), and generates an accessible interactive HTML report that enables clinician review. The integration of STRchive annotations provides immediate clinical context for findings at established disease loci, including pathogenic thresholds, inheritance patterns, and associated phenotypes.

## DATA AND CODE AVAILABILITY

The TRoLR workflow is available on GitHub (https://github.com/millerlaboratory/TRoLR) Full vamos genotypes, reference catalog data files, scripts and additional supplementary material is available on AWS (https://s3.amazonaws.com/1000g-ont/index.html?prefix=Gibson_etal_TRoLR_preprint_supplemental/). Some genotype and phenotypic data are available in dbGaP for UDN participants.

## ACKNOWLEDGEMENTS

We would like to acknowledge the Human Pangenome Reference Consortium (BioProject ID: PRJNA730823) and its funder, the National Human Genome Research Institute (NHGRI). This work was supported by NIH grants U01NS134355 and the UDN Data Management Coordinating Center (DMCC) U2CNS132415. HD is supported by NHGRI grant 4R00HG012796-03 and NHMRC Investigator grant GNT2026126. DEM is supported by NIH grant DP5OD033357.

## REFERENCES

1. Splinter, K. et al. Effect of genetic diagnosis on patients with previously undiagnosed disease. N. Engl. J. Med. 379, 2131–2139 (2018).

2. Porubsky, D. et al. Human de novo mutation rates from a four-generation pedigree reference. Nature 643, 427–436 (2025).

3. Steely, C. J., Watkins, W. S., Baird, L. & Jorde, L. B. The mutational dynamics of short tandem repeats in large, multigenerational families. Genome Biol. 23, 253 (2022).

4. Depienne, C. & Mandel, J.-L. 30 years of repeat expansion disorders: What have we learned and what are the remaining challenges? Am. J. Hum. Genet. 108, 764–785 (2021).

5. Paulson, H. Repeat expansion diseases. Handb. Clin. Neurol. 147, 105–123 (2018).

6. Hiatt, L. et al. STRchive: a dynamic resource detailing population-level and locus-specific insights at tandem repeat disease loci. Genome Med. 17, 29 (2025).

7. Dolzhenko, E. et al. ExpansionHunter: a sequence-graph-based tool to analyze variation in short tandem repeat regions. Bioinformatics 35, 4754–4756 (2019).

8. Dashnow, H. et al. STRetch: detecting and discovering pathogenic short tandem repeat expansions. Genome Biol. 19, 121 (2018).

9. Dashnow, H. et al. STRling: a k-mer counting approach that detects short tandem repeat expansions at known and novel loci. Genome Biol. 23, 257 (2022).

10. Dolzhenko, E. et al. ExpansionHunter Denovo: a computational method for locating known and novel repeat expansions in short-read sequencing data. Genome Biol. 21, 102 (2020).

11. Mousavi, N., Shleizer-Burko, S., Yanicky, R. & Gymrek, M. Profiling the genome-wide landscape of tandem repeat expansions. Nucleic Acids Res. 47, e90 (2019).

12. Gustafson, J. A. et al. High-coverage nanopore sequencing of samples from the 1000 Genomes Project to build a comprehensive catalog of human genetic variation. Genome Res. gr.279273.124 (2024).

13. Logsdon, G. A., Vollger, M. R. & Eichler, E. E. Long-read human genome sequencing and its applications. Nat. Rev. Genet. 21, 597–614 (2020).

14. Mantere, T., Kersten, S. & Hoischen, A. Long-read sequencing emerging in medical genetics. Front. Genet. 10, 426 (2019).

15. Miller, D. E. et al. Targeted long-read sequencing identifies missing disease-causing variation. Am. J. Hum. Genet. 108, 1436–1449 (2021).

16. Hiatt, S. M. et al. Long-read genome sequencing for the molecular diagnosis of neurodevelopmental disorders. HGG Adv. 2, 100023 (2021).

17. Sanford Kobayashi, E., et al. Approaches to long-read sequencing in a clinical setting to improve diagnostic rate. Sci. Rep. 12, 16945 (2022).

18. Ren, J., Gu, B. & Chaisson, M. J. P. vamos: variable-number tandem repeats annotation using efficient motif sets. Genome Biol. 24, 175 (2023).

19. Dolzhenko, E. et al. Characterization and visualization of tandem repeats at genome scale. Nat. Biotechnol. 1–9 (2024).

20. Chiu, R., Rajan-Babu, I.-S., Friedman, J. M. & Birol, I. Straglr: discovering and genotyping tandem repeat expansions using whole genome long-read sequences. Genome Biol. 22, 224 (2021).

21. Lougheed, D. R., Pastinen, T. & Bourque, G. STRkit: precise, read-level genotyping of short tandem repeats using long reads and single-nucleotide variation. bioRxiv 2025.03.25.645269 (2025) doi:10.1101/2025.03.25.645269.

22. Tesi, N. et al. Characterizing tandem repeat complexities across long-read sequencing platforms with TREAT and otter. Genome Res. 34, 1942–1953 (2024).

23. Danzi, M. C. et al. Detailed tandem repeat allele profiling in 1,027 long-read genomes reveals genome-wide patterns of pathogenicity. bioRxivorg 2025.01.06.631535 (2025) doi:10.1101/2025.01.06.631535.

24. Garimella, K. V. et al. Population-scale long-read sequencing in the All of Us Research Program. medRxiv 2025.10.02.25336942 (2025) doi:10.1101/2025.10.02.25336942.

25. Genetic Modifiers of Huntington’s Disease (GeM-HD) Consortium. Electronic address: & Genetic Modifiers of Huntington’s Disease (GeM-HD) Consortium. CAG repeat not polyglutamine length determines timing of Huntington’s disease onset. Cell 178, 887–900.e14 (2019).

26. Liao, W.-W. et al. A draft human pangenome reference. Nature 617, 312–324 (2023).

27. Ebert, P. et al. Haplotype-resolved diverse human genomes and integrated analysis of structural variation. Science 372, eabf7117 (2021).

28. Schloissnig, S. et al. Structural variation in 1,019 diverse humans based on long-read sequencing. Nature 1–11 (2025).

29. Weisburd, B. et al. Defining a tandem repeat catalog and variation clusters for genome-wide analyses. bioRxivorg 2024.10.04.615514 (2025) doi:10.1101/2024.10.04.615514.

30. Cui, Y. et al. A genome-wide spectrum of tandem repeat expansions in 338,963 humans. Cell 187, 2336–2341.e5 (2024).

31. Ramoni, R. B. et al. The Undiagnosed Diseases Network: Accelerating discovery about Health and disease. Am. J. Hum. Genet. 100, 185–192 (2017).

32. Cheng, H. et al. Efficient near-telomere-to-telomere assembly of nanopore simplex reads. Nature 1–8 (2026).

33. Danecek, P. et al. Twelve years of SAMtools and BCFtools. Gigascience 10, (2021).

34. Mikheenko, A., Prjibelski, A., Saveliev, V., Antipov, D. & Gurevich, A. Versatile genome assembly evaluation with QUAST-LG. Bioinformatics 34, i142–i150 (2018).

35. Gu, B. & Chaisson, M. TRcompDB v2.0, a global reference of human tandem repeat composition and mutation rate from long-read assemblies. Zenodo 10.5281/ZENODO.16286509 (2026).

36. Stevanovski, I. et al. Comprehensive genetic diagnosis of tandem repeat expansion disorders with programmable targeted nanopore sequencing. Sci. Adv. 8, eabm5386 (2022).

37. McDaniel, D. O., Keats, B., Vedanarayanan, V. V. & Subramony, S. H. Sequence variation in GAA repeat expansions may cause differential phenotype display in Friedreich’s ataxia. Mov. Disord. 16, 1153–1158 (2001).

38. Moseley, M. L. et al. Bidirectional expression of CUG and CAG expansion transcripts and intranuclear polyglutamine inclusions in spinocerebellar ataxia type 8. Nat. Genet. 38, 758–769 (2006).

39. Cleary, J. D., Subramony, S. H. & Ranum, L. P. W. Spinocerebellar ataxia type 8. In GeneReviews(®) (University of Washington, Seattle, Seattle (WA), 1993).

40. Chen, X. et al. Apolipoprotein E epsilon4 allele is associated with better performance language and visual memory in spinocerebellar ataxia type 3. Eur. J. Neurol. 32, e70017 (2025).

41. Tan, D. et al. CAG repeat expansion in THAP11 is associated with a novel spinocerebellar ataxia. Mov. Disord. 38, 1282–1293 (2023).

42. Zou, J. et al. A Chinese SCA36 pedigree analysis of NOP56 expansion region based on long-read sequencing. Front. Genet. 14, 1110307 (2023).

43. Hunter, J. E., Berry-Kravis, E., Hipp, H. & Todd, P. K. FMR1 disorders. In GeneReviews(®) (University of Washington, Seattle, Seattle (WA), 1993).

44. Liu, T. et al. Simultaneous screening of the FRAXA and FRAXE loci for rapid detection of FMR1 CGG and/or AFF2 CCG repeat expansions by triplet-primed PCR. J. Mol. Diagn. 23, 941–951 (2021).

45. Rasmussen, A. et al. High resolution analysis of DMPK hypermethylation and repeat interruptions in myotonic dystrophy type 1. Genes (Basel) 13, 970 (2022).

46. Kurosaki, T. & Ashizawa, T. The genetic and molecular features of the intronic pentanucleotide repeat expansion in spinocerebellar ataxia type 10. Front. Genet. 13, 936869 (2022).

47. Ardui, S. et al. Detecting AGG interruptions in females with a FMR1 premutation by long-read single-molecule sequencing: A 1 year clinical experience. Front. Genet. 9, 150 (2018).

48. Akçimen, F. et al. Long-read sequencing identifies FGF14 repeat expansions in Parkinson’s disease. Brain (2025) doi:10.1093/brain/awaf456.

49. Zheng, X. et al. A complex FGF14 (TTC)/(TGC) repeat expansion in Parkinson’s disease. Mov. Disord. 41, 667–678 (2026).

50. Ishiura, H. et al. Noncoding CGG repeat expansions in neuronal intranuclear inclusion disease, oculopharyngodistal myopathy and an overlapping disease. Nat. Genet. 51, 1222–1232 (2019).

51. Granhøj, J. et al. MUC1-associated autosomal dominant tubulointerstitial kidney disease: prevalence in kidney failure of undetermined aetiology and clinical insights from Danish families. Clin. Kidney J. 18, sfae355 (2025).

52. Kirby, A. et al. Mutations causing medullary cystic kidney disease type 1 lie in a large VNTR in MUC1 missed by massively parallel sequencing. Nat. Genet. 45, 299–303 (2013).

53. Chen, S. et al. A genomic mutational constraint map using variation in 76,156 human genomes. Nature 625, 92–100 (2024).

54. Xu, S. et al. Use ggbreak to Effectively Utilize Plotting Space to Deal With Large Datasets and Outliers. Front. Genet. 12, 774846 (2021).

55. Readman, C., Indhu-Shree, R.-B., Jan M, F. & Inanc, B. A comprehensive tandem repeat catalog of the human genome. Nat. Commun. 17, 1106 (2026).

56. 1000 Genomes Project Consortium et al. A global reference for human genetic variation. Nature 526, 68–74 (2015).

57. Jensen, T. D. et al. Integration of transcriptomics and long-read genomics prioritizes structural variants in rare disease. Genome Res. 35, 914–928 (2025).

58. LaFlamme, C. W. et al. Diagnostic utility of DNA methylation analysis in genetically unsolved pediatric epilepsies and CHD2 episignature refinement. Nat. Commun. 15, 6524 (2024).

59. Akçimen, F. et al. Expanded CAG repeats in ATXN1, ATXN2, ATXN3, and HTT in the 1000 Genomes Project. Mov. Disord. 36, 514–518 (2021).

60. Wallace, S. E. & Bean, L. J. H. Resources for Genetics Professionals — Genetic Disorders Caused by Nucleotide Repeat Expansions and Contractions. (University of Washington, Seattle, 2022).

61. Vegezzi, E. et al. Neurological disorders caused by novel non-coding repeat expansions: clinical features and differential diagnosis. Lancet Neurol. 23, 725–739 (2024).

62. Deng, J. et al. Expansion of GGC repeat in GIPC1 is associated with oculopharyngodistal myopathy. Am. J. Hum. Genet. 106, 793–804 (2020).

63. Boivin, M. et al. GGC repeat expansions within new open reading frames are translated into toxic polyglycine proteins in oculopharyngodistal myopathy. Nat. Genet. 1–13 (2026).

64. Marcogliese, P. C. et al. IRF2BPL is associated with neurological phenotypes. Am. J. Hum. Genet. 103, 245–260 (2018).

65. Venkateswaran, S. et al. IRF2BPL-related disorder, causing neurodevelopmental disorder with regression, abnormal movements, loss of speech and seizures (NEDAMSS) is characterized by pathology consistent with DRPLA. Mov. Disord. 39, 2102–2109 (2024).

66. Grube, S. et al. A CAG repeat polymorphism of KCNN3 predicts SK3 channel function and cognitive performance in schizophrenia: Genetic SK3 variation influences cognition. EMBO Mol. Med. 3, 309–319 (2011).

67. Ivković, M. et al. Schizophrenia and polymorphic CAG repeats array of calcium-activated potassium channel (KCNN3) gene in Serbian population. Int. J. Neurosci. 116, 157–164 (2006).

68. Padilla, L. et al. In silico prediction of the impact of genomic variations in the small conductance calcium activated potassium channel SK3 structure and function. Front. Neurosci. 19, 1631536 (2025).

69. Gatchel, J. R. & Zoghbi, H. Y. Diseases of unstable repeat expansion: mechanisms and common principles. Nat. Rev. Genet. 6, 743–755 (2005).

70. LaCroix, A. J. et al. GGC repeat expansion and Exon 1 methylation of XYLT1 is a common pathogenic variant in baratela-Scott syndrome. Am. J. Hum. Genet. 104, 35–44 (2019).

71. Sarafidou, T. et al. Folate-sensitive fragile site FRA10A is due to an expansion of a CGG repeat in a novel gene, FRA10AC1, encoding a nuclear protein. Genomics 84, 69–81 (2004).

72. Van de Vondel, L. et al. A 5’ UTR CCG expansion in TBC1D7 causes oculopharyngodistal myopathy. medRxiv 2026.03.27.26349107 (2026) doi:10.64898/2026.03.27.26349107.

73. Fazal, S. et al. A genome-wide approach for the discovery of novel repeat expansion disorders in the Undiagnosed Diseases Network cohort. Genet. Med. 27, 101462 (2025).

74. Wieben, E. D. et al. Comprehensive assessment of genetic variants within TCF4 in Fuchs’ endothelial corneal dystrophy. Invest. Ophthalmol. Vis. Sci. 55, 6101–6107 (2014).

75. Butland, S. L. et al. CAG-encoded polyglutamine length polymorphism in the human genome. BMC Genomics 8, 126 (2007).

76. Delahaye, C. & Nicolas, J. Sequencing DNA with nanopores: Troubles and biases. PLoS One 16, e0257521 (2021).

77. Park, G., An, H., Luo, H. & Park, J. NanoMnT: an STR analysis tool for Oxford Nanopore sequencing data driven by a comprehensive analysis of error profile in STR regions. Gigascience 14, giaf013 (2025).

