## Supplemental Figures for "Genome-wide detection and clinical prioritization of tandem repeat outliers using long-read sequencing"

**TRoLR Supplemental Figures**

- **Figure S1: Example TRoLR interactive HTML report**
- **Figure S2: Comparison of 99th percentile LPS values to established pathogenic thresholds across STRchive loci**
- **Figure S3: Expanded alleles at autosomal dominant STRchive loci identified in the reference cohort**
- **Figure S4: Expanded alleles at autosomal recessive and X-linked STRchive loci in the reference cohort**
- **Figure S5: Expanded alleles with benign motifs at STRchive loci associated with autosomal dominant conditions**
- **Figure S6: GIPC1 GGC repeat expansion with associated hypermethylation in a control cohort individual**
- **Figure S7: CGG repeat expansion outliers without hypermethylation**
- **Figure S8:** IGV screenshots of positive controls

**Figure S1**

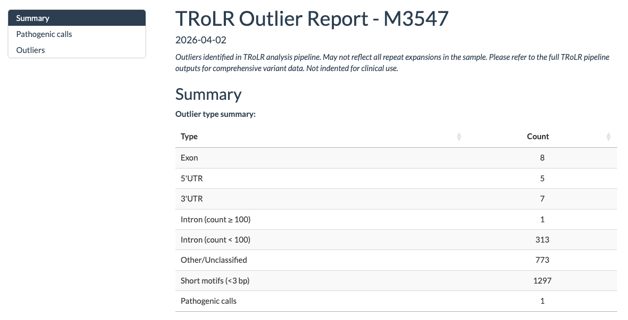

**A**

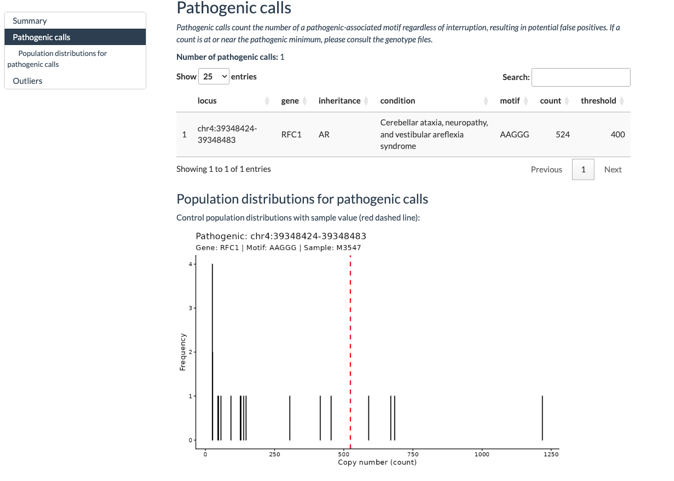

**B**

**Figure S1**

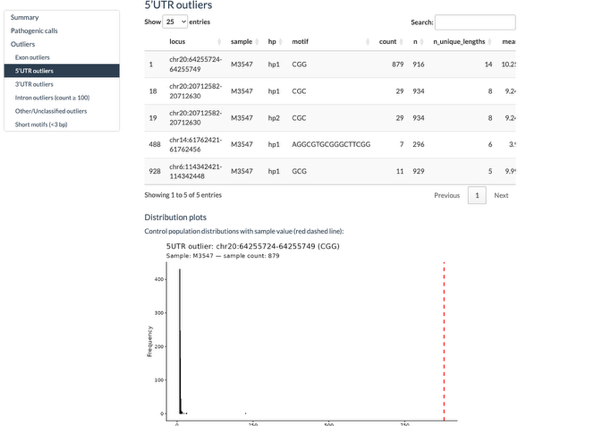

**C**

**Figure S1: Example TRoLR interactive HTML report.** Screenshots from the TRoLR output report for a representative UDN sample (M3547). **(A)** Summary page showing the number of outlier loci identified, broken down by genomic context (exon, 5'UTR, 3'UTR, intronic ≥100 copies, intronic <100 copies, other/unclassified, and short motifs <3 bp), along with the number of STRchive pathogenic calls. **(B)** Pathogenic calls page displaying the RFC1 AAGGG expansion identified in this individual (524 repeat units, pathogenic threshold 400 units), with an embedded histogram showing the population distribution of repeat counts at this locus. The red dashed line indicates the sample's value. **(C)** Outlier table for 5'UTR loci, showing the PCMTD2 CGG expansion (879 repeat units) as the top-ranked outlier. An embedded distribution plot displays the population LPS distribution with the sample value indicated.

**Figure S2**

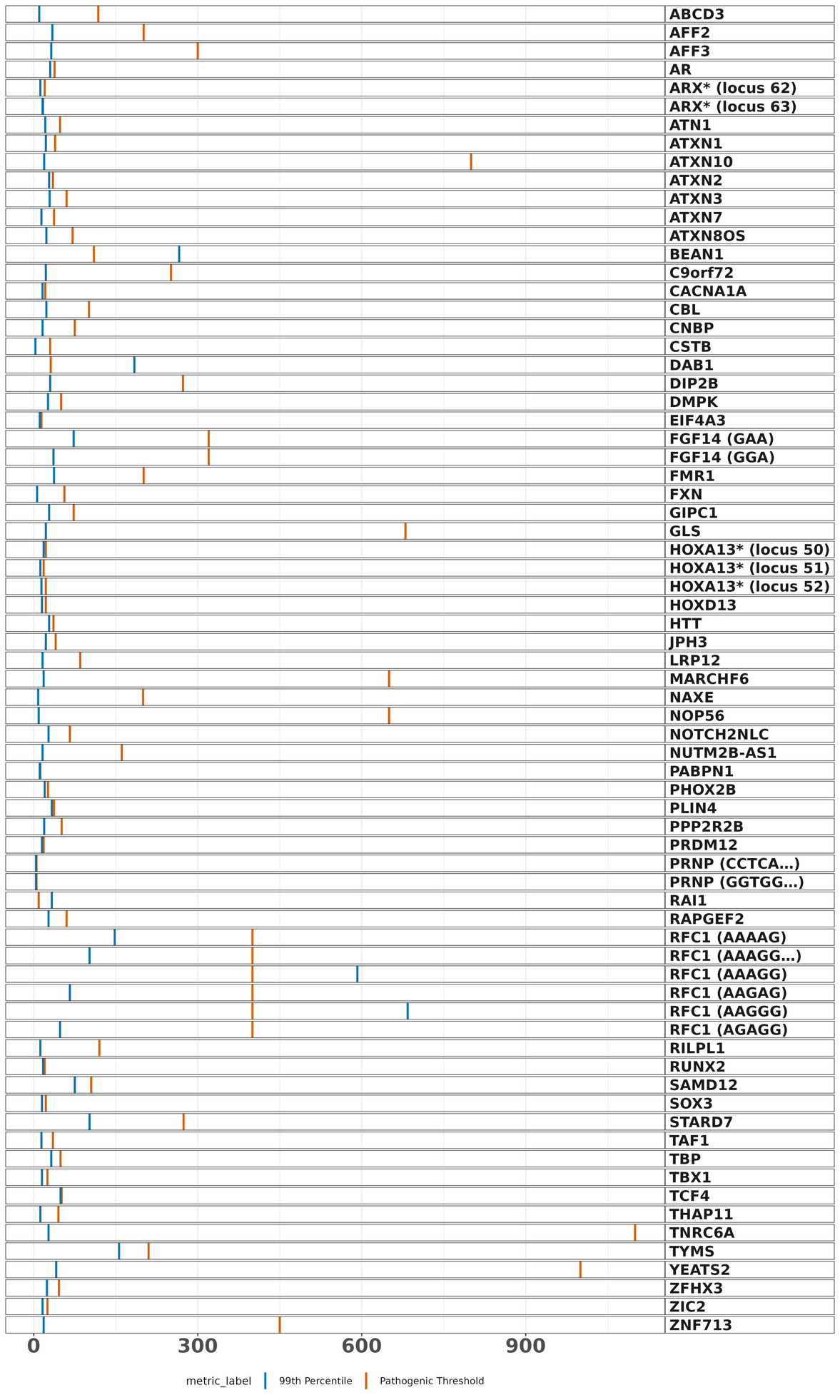

**Figure S2: Comparison of 99th percentile LPS values to established pathogenic thresholds across STRchive loci.** For each of the 65 STRchive loci genotyped and included in reports, the 99th percentile LPS from the population distribution (blue) is shown alongside the established pathogenic minimum threshold (orange). At most loci, the 99th percentile falls below the pathogenic minimum, confirming that the 99th percentile threshold does not generate false positive pathogenic calls in the reference cohort. Loci are ordered alphabetically by gene name. Labels with an asterisk and a locus number identifier are two separate loci in the same gene. Labels with a motif in parentheses are loci with multiple LPS motifs at the same locus.

**Figure S3**

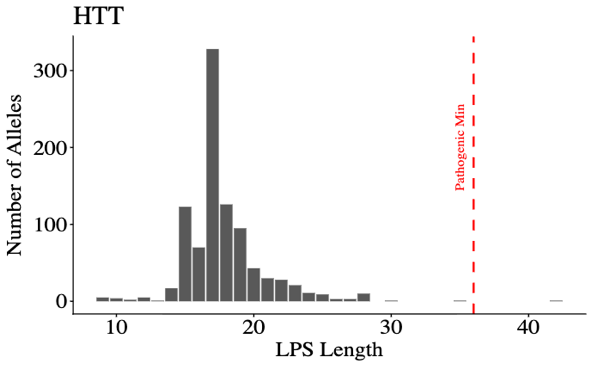

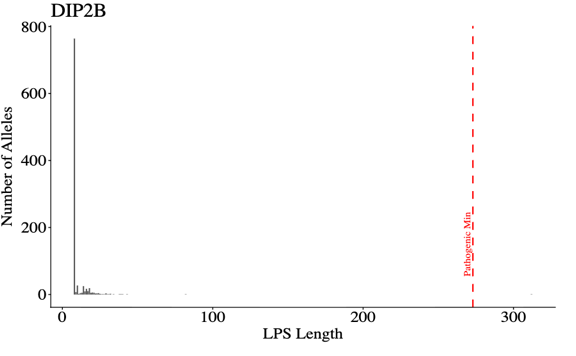

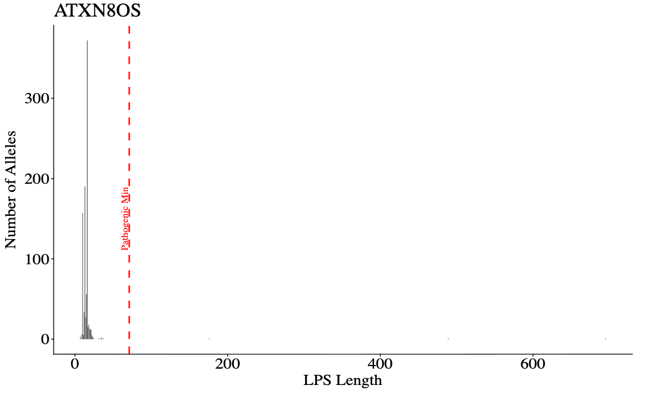

**A**

**B**

**C**

**Figure S3: Expanded alleles at autosomal dominant STRchive loci identified in the reference cohort.** Histograms of LPS distributions for **(A)** HTT, **(B)** DIP2B, and **(C)** ATXN8OS. Red dashed lines indicate established pathogenic thresholds. In HTT, one individual (HG02470) carried an allele of 42 CAG repeats, exceeding the pathogenic threshold of 39 units. In DIP2B, one individual (NA20752) carried an expanded allele. In ATXN8OS, three individuals (HG02055, HG02258, HG02514) carried expanded alleles exceeding the pathogenic threshold. These individuals are from the 1000 Genomes Project and phenotypic information is not available.

**Figure S4**

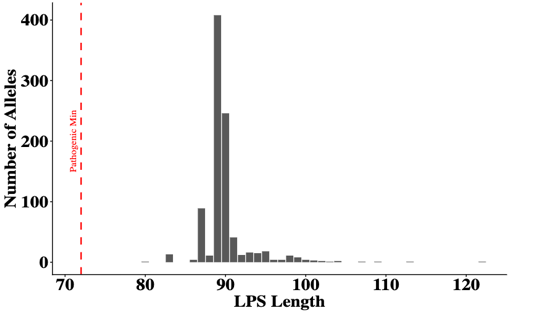

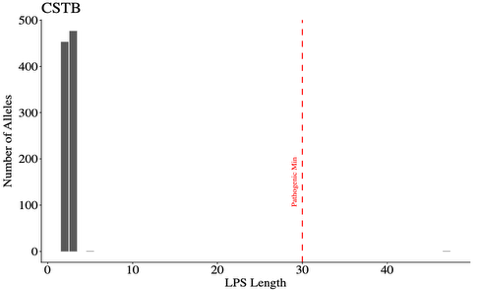

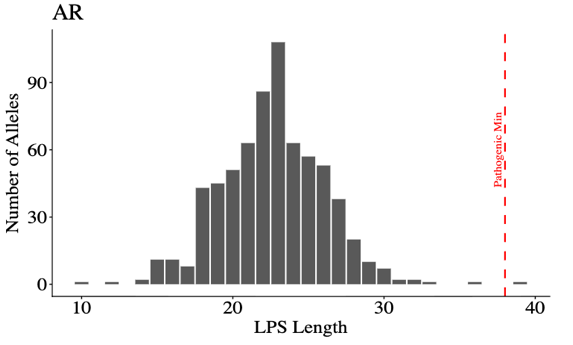

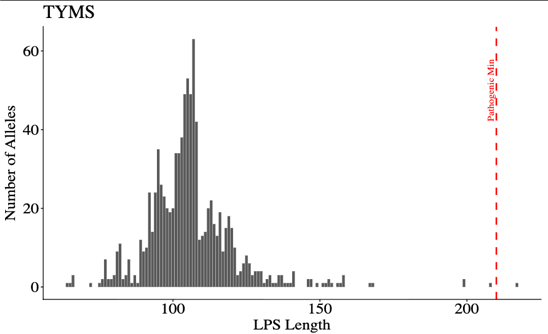

**A**

**B**

**C**

**D**

**Figure S4: Expanded alleles at autosomal recessive and X-linked STRchive loci in the reference cohort.** Histograms of LPS distributions for **(A)** CSTB, **(B)** AR, **(C)** TYMS, **(D)** XYLT1. Red dashed lines indicate established pathogenic thresholds. One carrier allele was identified at *CSTB*, *AR*, and *TYMS* locus, consistent with expected population carrier frequencies for CSTB and AR. All *XYLT1* loci are over the pathogenic threshold, but without the associated hypermethylation.

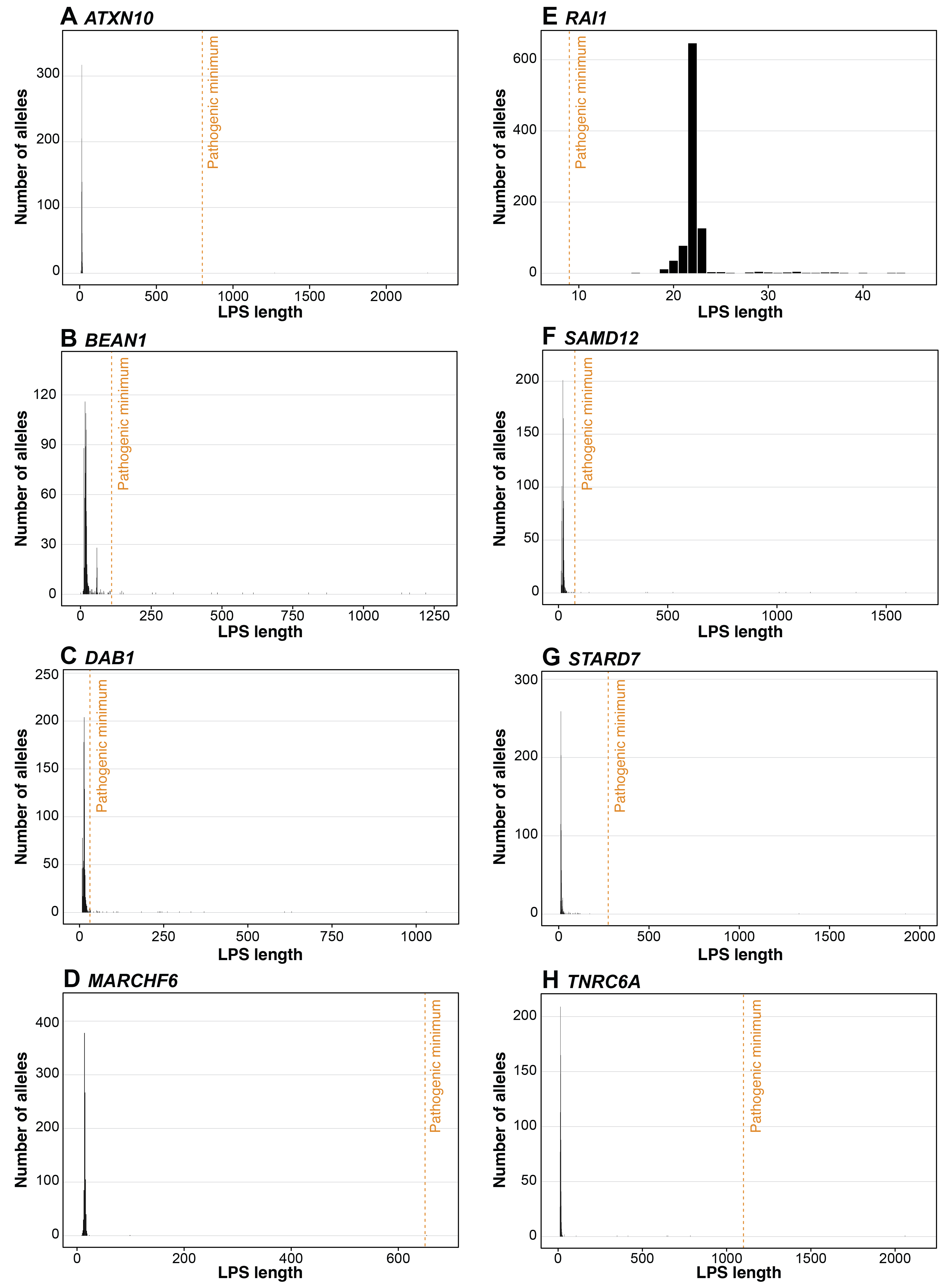
**Figure S5**

**Figure S5: Expanded alleles with benign motifs at STRchive loci associated with autosomal dominant conditions.** **(A)** Histogram of LPS distribution at ATXN10, showing multiple individuals with expansions exceeding the pathogenic threshold but composed of the benign ATTCT motif rather than the pathogenic ATTCC motif. **(B)** Histogram of LPS distribution BEAN1, showing multiple individuals with expansions exceeding the pathogenic threshold but composed of the benign AATAA motif rather than the pathogenic TGGAA motif. **(C)** Histogram of LPS distribution at DAB1, showing multiple individuals with expansions exceeding the pathogenic threshold but composed of the benign AAAAT motif rather than the pathogenic GAAAT motif. **(D)** Histogram of LPS distribution at MARCHF6, showing one individual with expansions exceeding the pathogenic threshold but composed of the benign TTTTA motif rather than the pathogenic TTTCA motif **(E)** Histogram of LPS distribution at RAI1, showing multiple individuals with expansions exceeding the pathogenic threshold but composed of the benign TTTTA motif rather than the pathogenic TTTCA motif **(F)** Histogram of LPS distribution at SAMD12, showing multiple individuals with expansions exceeding the pathogenic threshold but composed of the benign TAAAA motif rather than the pathogenic TGAAA motif **(G)** Histogram of LPS distribution at STARD7, showing multiple individuals with expansions exceeding the pathogenic threshold but composed of the benign AAAAT motif rather than the pathogenic AAATG motif **(H)** Histogram of LPS distribution at TNRCA, showing multiple individuals with expansions exceeding the pathogenic threshold but composed of the benign TTTTA motif rather than the pathogenic TTTCA motif.

**Figure S6**

**A**

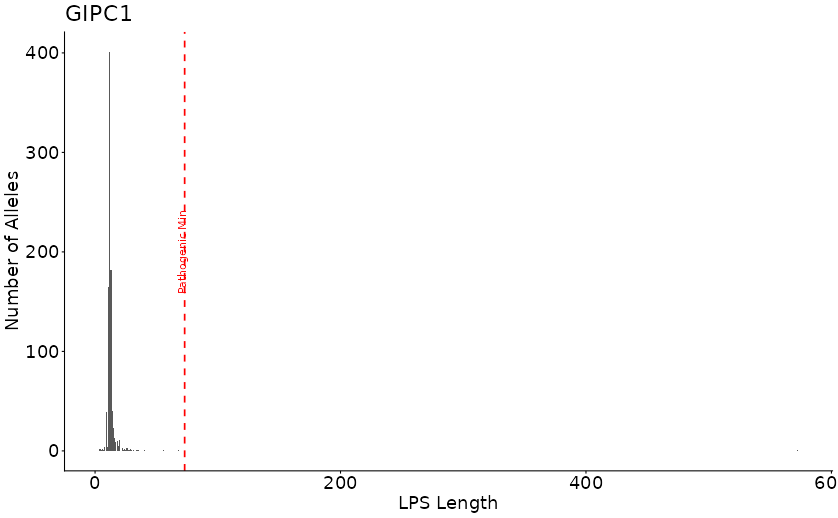

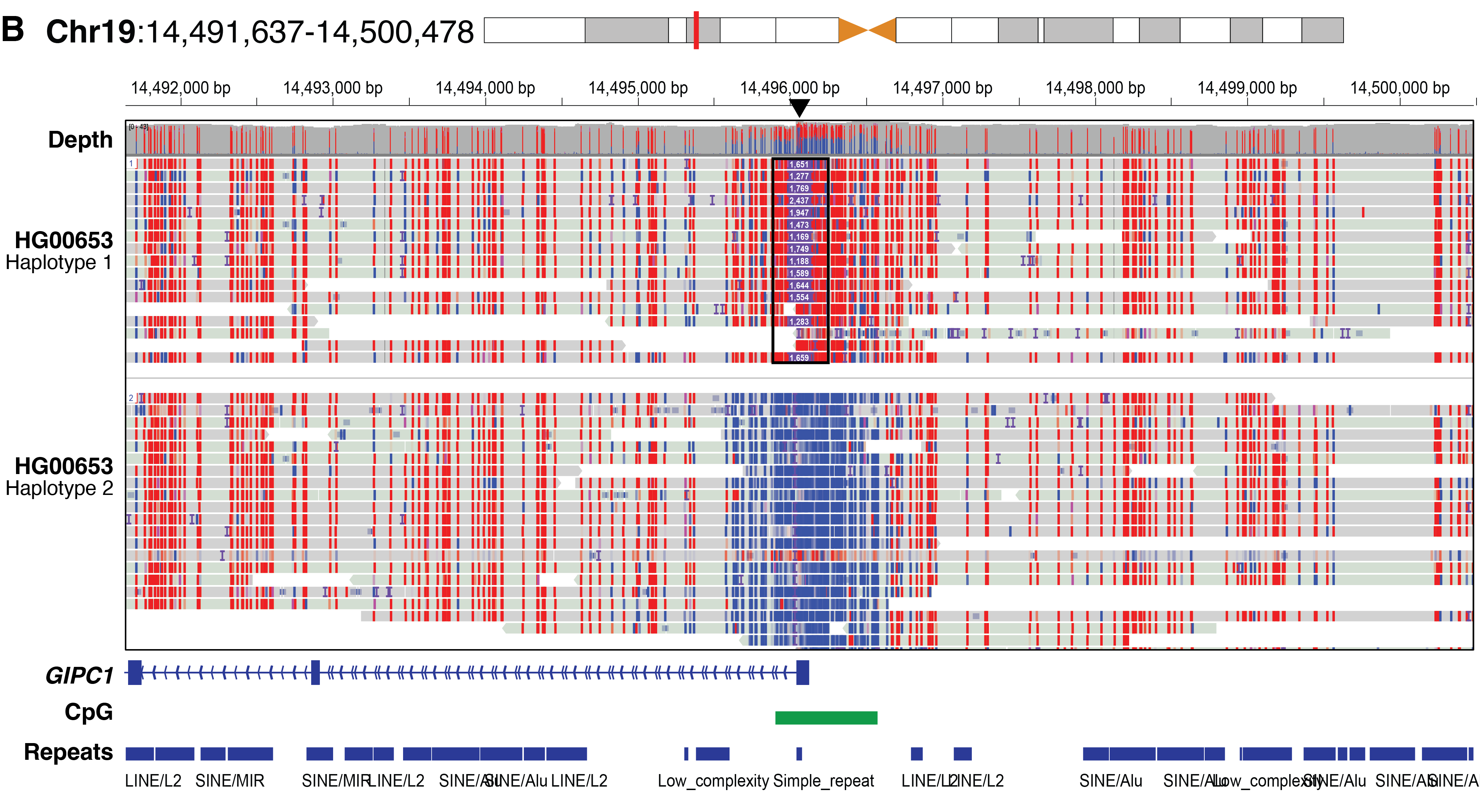

**Figure S6: *GIPC1* GGC repeat expansion with associated hypermethylation in a control cohort individual.** **(A)** Histogram of LPS values at the GIPC1 locus across the reference cohort. The red dashed line indicates the established pathogenic threshold (73 repeats). One individual (HG00653, East Asian ancestry) carried an expansion of 571 GGC units, approximately fivefold larger than the pathogenic range. **(B)** IGV visualization of nanopore reads at the GIPC1 locus in HG00653, colored by 5-methylcytosine probability, demonstrating hypermethylation of the expanded allele. Expansions at this locus cause oculopharyngodistal myopathy type 2 (OPDM2).

**Figure S7**

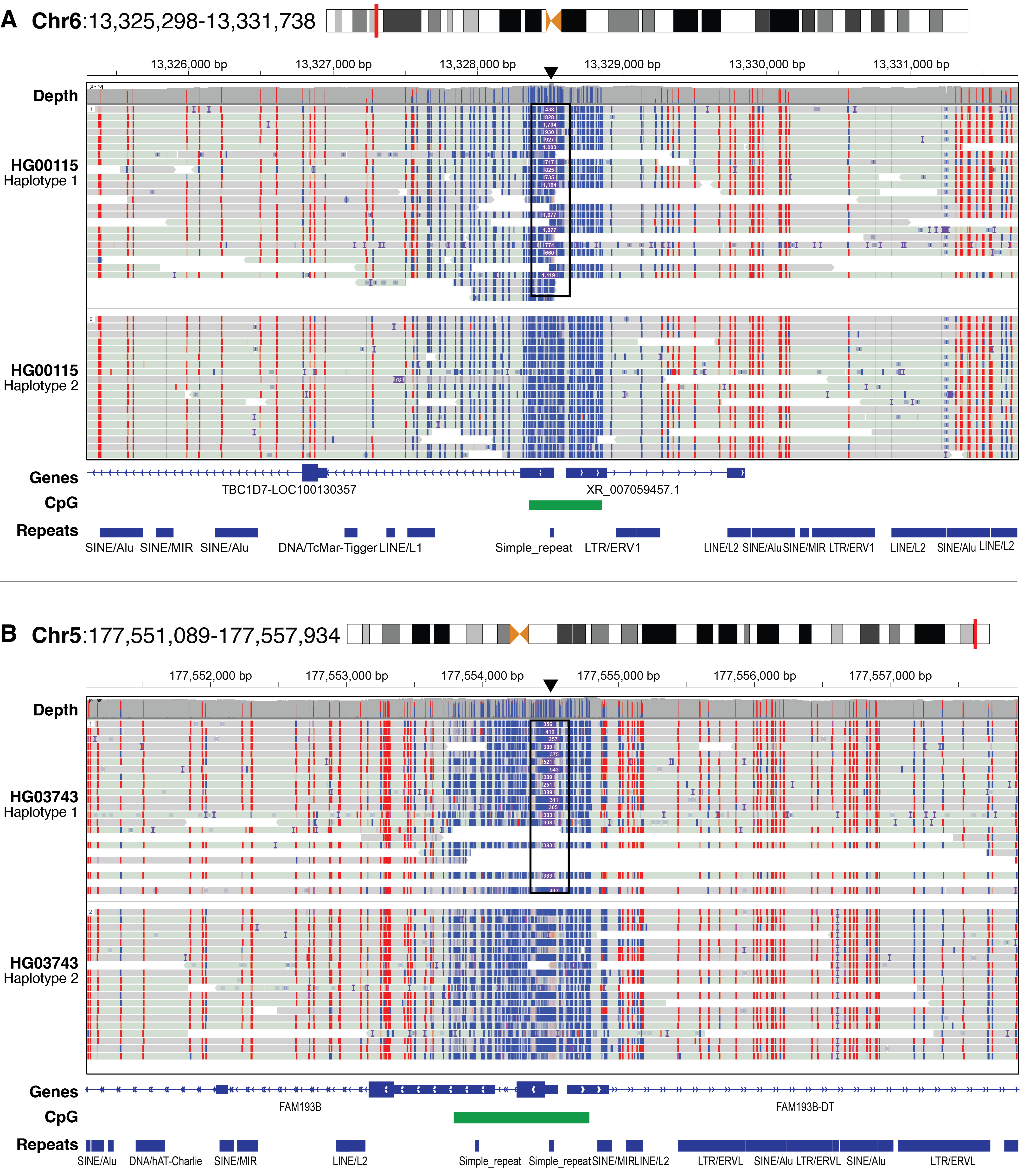

**Figure S7: CGG repeat expansion outliers without hypermethylation.** IGV visualization of nanopore reads at **(A) the** TBC1D7 expansion (HG00115, ~311 CGG repeats) and **(B)** the *FAM193B* (HG03743, 147 repeats)*.* Expansions at these loci are suspected to cause OPMD.

**Figure S8**

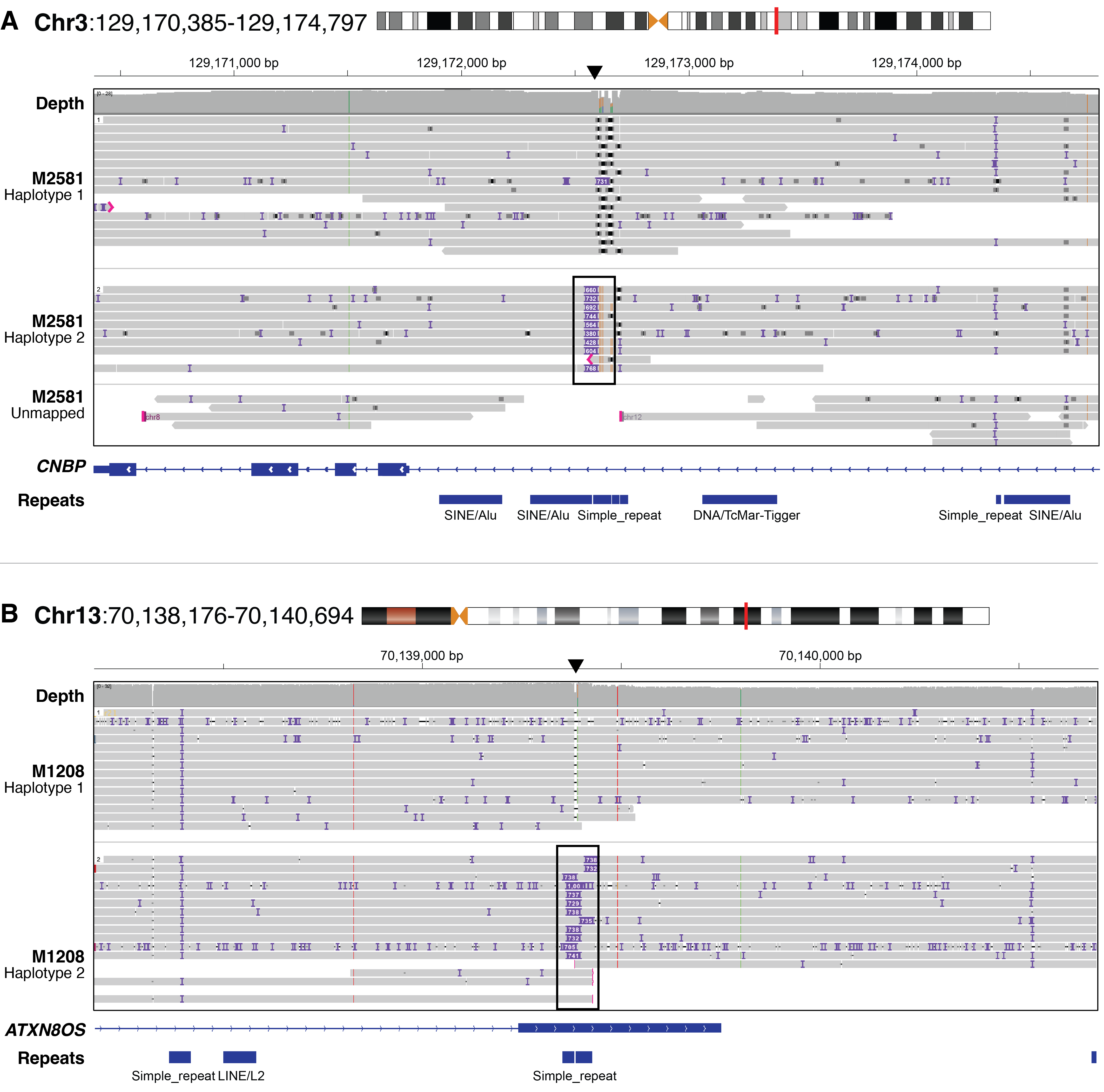

**Figure S8: Repeat expansions identified in 2 UDN cases used as positive controls.** IGV view of reads at the **(A)** *CNBP* locus, which was confirmed by clinical testing, and **(B)** at the *ATXN8OS* locus, which was confirmed by PCR.
